# The Social Sunshine of the Arctic Youth: Exploring friendship’s influence on Vitamin D levels

**DOI:** 10.1101/2023.11.29.23299188

**Authors:** Rafael A. Nozal Cañadas, Christopher Sivert Nielsen, Anne-Sofie Furberg, Anne Merethe Hanssen, Lars Ailo Bongo

## Abstract

**Background:** Vitamin D status correlates with 25OHD levels which depends on nutritional intake and UVB exposure. These two factors are influenced by friends, as people tend to participate in the same activities or eat a similar diet to their peers.

**Objectives:** Investigate how social interactions in a general high school population above the Arctic Circle influence 25OHD levels in the population.

**Methods:** The Fit Futures 1 study was performed over 8 months and interview data on social contact among 1038 first-level students in 8 high schools in Northern Norway were collected. Serum levels of 25OHD were measured (n = 890). The participants filled in a questionnaire about nutritional consumption, solarium habits, ethnicity, and chronic diseases. The participant’s BMI was also measured.

**Results:** Once high schools’ social biases were accounted for, only UVB radiation levels explained the differences in 25OHD levels. For non-solarium users, logistic regression analysis showed a positive correlation between a person’s 25OHD levels and the 25OHD level of friends in the general population and within the same high school for most of the schools. We saw that women can influence other women into going to the solarium, while this influence was not present for men.

**Conclusions:** 25OHD levels can be influenced by social networks. This study can help to add weight to current public health recommendations due to the positive spillover effect in the network.

## 1 Introduction

Humans tend to form friendship ties with individuals who are similar to them in certain aspects, referred to as homophily [1]. Although there is vast evidence that homophily shapes social networks, there is limited insight into how homophily functions in these networks. Most studies of homophily have been concerned with demographic variables such as age, sex, and social class. More recent evidence demonstrates deeper similarities among friends in behavior and personality [2]. Christiakis and Fowler showed a spread of obesity through social ties [3]. If friends are more similar to one another in terms of healthy or unhealthy behavior, then social network proximity should be associated with similarity in biomarker profiles. Whether or not adolescents tend to associate with others who have similar lifestyles and experience similar lifestyle-related biomarker profiles has yet to be evaluated.

Vitamin D deficiency is emerging as a very common condition worldwide [4] and is associated with unfavorable skeletal outcomes, excess mortality, and a higher risk of infections. Serum 25-hydroxyvitamin D (25OH)D is considered to be the best biomarker of the body’s vitamin D status and integrates Vitamin D derived from dietary intake and cutaneous synthesis after exposure to ultraviolet B (UVB) radiation of the solar electromagnetic spectrum. It has been estimated that the general European population gets 80-90% of its vitamin D from endogenous production in UVB-exposed skin. There is, however, considerable variation in this proportion across populations, population groups, and between individuals. Importantly, populations living at higher latitudes with periodic lack of photosynthesis may be at higher risk of vitamin D deficiency. Among a general youth population at 69°N participating in the Fit futures study, Tromsø Norway, 60% had vitamin D deficiency, defined by serum 25OHD below 50 nmol/l [5]. In the general adult population participating in the Tromsø Study, 19% had vitamin D deficiency, which is lower than the prevalence found in national data for Norway (28%) and in studies among adult populations further south [6–10].

Data from the Fit Futures and Tromsø study suggest that a sufficient level of serum 25(OH)D reflects several healthy lifestyle factors, such as outdoor activities, lower BMI, and fish-rich diet [5, 11], which may be defined by social interactions.

To our knowledge, no previous studies have been done on the effect of social networks in relation to vitamin D levels. Only one previous study has shown that poor economic factors influence health, because of the lack of vitamin D [12]. There has been a shift in social dynamics with indoor isolation during the COVID-19 pandemic, which showed how the severity of SARS-Covid-19 was linked to vitamin D deficiency [13]. Finally, ethnicity tends to be a strong social cohesion factor [14–26], and people of Middle Eastern, black, and South Asian sea descent require higher UVB levels and show higher deficiency prevalence than the white population [27–33]. Our objectives are to perform an explorative analysis in a general youth high school population to determine if social dynamics can affect 25OHD levels, and if so, to what extent environmental and lifestyle factors follow the same dynamics.

## 2 Methods

### 2.1 Population and study design

The Fit Futures (FF) study [34] is a cohort with repeated health surveys among students from 8 high schools (H1-H8) in the Norwegian municipalities of Tromsø and Balsfjord (supplementary figure 4). FF1 was conducted from October 2010 to May 2011 (supplementary figure 5). All first-year students in the 8 high schools were invited (supplementary table 3), with consecutive inclusion of the eight schools. A total of 1117 youths were invited 93% attended, 508 girls (48.9%), and 530 boys. The age ranges from 15 to 28 years old, with 822 (79.2%) being 16 years or younger, and 52 (5%) older than 18 years. Students with special educational needs or mental disabilities are allowed to study for several years in high school in Norway.

The participants had a one-day visit to The Clinical Research Unit at the University Hospital of North Norway (UNN Tromsø), which included clinical examinations, microbiological samples, blood samples, an interview (self-reported social network, acute and chronic disease, medication, pregnancy), and a web-based general questionnaire. All procedures were performed by trained research nurses.

### 2.2 Social network assessment

The social network was constructed based on the following question: *“Which students have you had the most contact with the last week? Name up to 5 students at your own school or other schools in Tromsø and Balsfjord.”*. Reciprocity in the nomination was not mandatory. For each of the nominations, five “yes/no” questions assessed the type of contact they had with their nominations: *“Do you have physical contact?”*, *“Are you together at school?”*, *“Are you together at sports?”*, *“Are you together at home?”*, *“Are you together at other places?”*. This resulted in five social networks: Physical, School, Sport, Home, and Other. Adding all the relationships together formed the Overall Network. Illustrations for each network are presented in the supplementary materials (supplementary figure 6).

To evaluate if the friends mentioned were representative of the participant’s social network, the following question was asked: *“To what degree does this table of friends give an overview of your social network? Please indicate on a scale from 0 (small degree) to 10 (high degree).”* (supplementary figure 7).

### 2.3 Vitamin D assessment

Non-fasting blood samples were collected from an antecubital vein, and serum was separated and frozen at −70°C in the Fit Futures Biobank at the UiT The Arctic University of Norway. All serum samples (n = 890) were sent to the Hormone Laboratory, Haukeland University Hospital, Bergen, Norway; and 25OHD, 25OHD2, and 25OHD3 were analyzed by high-pressure liquid chromatography-mass spectroscopy (LC-MS/MS). A sample from all blood vials was reanalyzed at University College Cork, Cork, Ireland, by LC-MS/MS again as a part of the Vitamin D Standardization Project (VDSP) [35], and standardization was applied to the rest of the samples [36].

25OHD was used as a marker for vitamin D levels. This combines both sources of provitamin D + UVB, and D2+D3 from diet. It has a longer half-life span in blood than other available metabolites.

In the analyses where categorical data is necessary, levels were defined as a binary variable dividing into vitamin D deficiency (< 50 nmol/L) or not vitamin D deficiency (>= 50 nmol/L). In all cases, vitamin D toxicity is defined as greater than 150 nmol/L. [37–40]

25OHD levels are increased during pregnancy. All women who reported a possible pregnancy were given pregnancy tests, which all came back negative. The skin also loses the efficiency of synthesizing vitamin D with age [41], since the population is composed of young adults, no further analysis taking age into consideration was performed.

### 2.4 Diseases and medicine usage

There is a low prevalence of vitamin D absorption-impairing diseases, which are also heterogeneously distributed among the study population. Nobody in our population reported having bariatric surgery.

We have no significant number of students taking vitamin D-influencing medication such as anti-seizure drugs [42–44], steroid drugs [45–51], fat absorption reduction [45, 52–54], cholesterol metabolism modification [45, 55–57] or diuretics. The only anti-inflammatories drug [45–51], reported is “Ibuprofen 200mg” (n = 120), which does not affect the vitamin D level.

### 2.5 Melanin levels and ethnicity classification

The participants answered the question *“Do you consider yourself as…”*, with possible answers *“Norwegian?”*, *“Sami”*, *“Kven / Finnish?”* or *“Other? (Please specify)”*. Also, two questions regarding the country of birth of parents: *“Was your biological mother/father born in Norway? If not specify”*. These questions are summarized into a single variable with the ethnicity, or combinations of ethnicities, for each participant (supplementary table 6).

We combined all answers (n = 1018) into a single melanin quantity binary variable, dividing for assumed *“Fair Skin”* (996) for those of European, North American, or North Asia background, or mixing of any of these; and *“Dark Skin”* (22) for anyone with South American, African, South Asian, or any mixed background that included these. No conditions related to albinism or hyperpigmentation were self-reported in this population.

### 2.6 Solarium assessment

Visits to the solarium were recorded by the question *“Have you used a solarium during the last 4 weeks?”*. In Norway, access to solarium was restricted for teenagers from 2012, but not law-enforced until 2017 [58]. This data was gathered during 2010 and 2011 before any restrictions. Students were divided into solarium and non-solarium users according to their answers.

### 2.7 Nutritional information

There were 6 relevant questions regarding vitamin D intake. *“How often do you usually eat fat fish (e.g. salmon, trout, mackerel, herring)”*, *“How often do you usually eat lean fish (e.g. cod, saithe, haddock)”*, *“How much do you usually drink of whole milk, kefir and yoghurt?”*, *“How often do you usually eat cheese (all kinds)?”*, *“Do you take cod liver oil, cod liver oil capsules or fish oil capsules?”*, *“Do you use vitamin or mineral supplements?”*. All answers were classified as categorical variables, ranging from never to every day.

In Norway at the time of the study, fortification of food with vitamin D was only common in low-fat milk and flavored milk [59] (0.4 µg vitamin D per 100g) and for baby formula (0.48–0.72 µg/100 kJ) [60].

Cholesterol is an important precursor in the transformation of 7-dehydro cholesterol (pro-vitamin D3) into pre-vitamin D3 via UVB catalyzation. Our population shows overwhelmingly healthy levels of HLD, and LDL, and no significant number of diseases that might affect the liver cholesterol biosynthetic pathway.

### 2.8 Anthropometric assessment

Weight and height were measured using an automatic electronic scale (Jenix DS 102 stadiometer, Dong Sahn Jenix, Seoul, Korea) with participants wearing light clothing and no footwear. Body mass index (BMI) is calculated as weight (kg) divided by the squared height (m2) with no correction for sex or age.

### 2.9 Physical activity assessment

The participants stated their physical activity level according to four hierarchical levels using a slightly modified version of the Saltin-Grimby Physical Activity Level Scale [61].

*“Exercise and physical exertion in leisure time. If your activity varies much, for example between summer and winter, then give an average”* with possible answers *“Reading, watching TV, or other sedentary activity?”*, *“Walking, cycling, or other forms of exercise at least 4 hours a week? (including walking or cycling to place of school, shopping, Sunday-walking, etc.)”*, *“Participation in recreational sports, heavy outdoor activities, snow clearing etc? (note: duration of activity at least 4 hours a week)”* and *“Participation in hard training or sports competitions, regularly several times a week?”*. The answers are shortened into “None”, “Light”, “Medium” and “Hard” respectively for convenience.

A related question was *“How many hours per day do you spend by the PC, watch TV, DVD etc. outside school during weekends?”* with answers ranging from *“None”* to *“10 hours or more”*.

### 2.10 Recreational drugs assessment

The use of recreational drugs was self-reported via the web-based questionnaire. For alcohol consumption, the question was *“How often do you drink alcohol?”*, with possible answers *“Never”*, *“Once per month or less”*, *“2-4 times per month”*, *“2-3 times per week”*, *“4 or more times per week”*. Due to the low number of answers in some of the categories, *“2-4 times per month”*, *“2-3 times per week”*, *“4 or more times per week”* are all combined into *“Twice per month or more”*. For smoking the question was *“Do you smoke?”* with possible answers *“No, never”*, *“Sometimes”* and *“Daily”*. For snuff use the question was *“Do you use snuff?”* with possible answers *“No, never”*, *“Sometimes”* and *“Daily”*.

### 2.11 Natural UVB light, sun irradiance, and polar night

Tromsø and Balsfjord are located inside the Artic Circle (60°+ N). The polar night started on the 23rd of November of 2010 and ended on the 19th of January 2011. During this time there is no sun irradiance. Solar irradiance above 60°N for 2011 is estimated to be about 8000 Wh/m² at its peak in July [62] (32.000 Wh/m² in the tropics at the same time [63]). After the polar night until the end of April, snow covers the ground; snow reflects UVB with about 86% efficiency. To determine which dates in our timeframe have relevant UVB, we estimated the duration of sun exposure to get 1000 IU of vitamin D synthesis. Assuming a clear day, snowy ground, type II skin, and 10% body exposure [64]: On March 1st it is not possible to acquire that amount. On March 15th it takes 5.22h. April 1st it takes 1.63h. On May 1st it takes 0.58h long. During autumn, assuming grass instead of snow, it is also not possible to acquire such an IU amount from October 1st.

Possible traveling was self-reported with the question: *“Have you been on a beach holiday during the last two months?”*, with only possible answers *“Yes”* and *“No”*. Traditionally, traveling might occur before high schools start near the end of August, in the nearly two weeks of winter break during Christmas, and a week during Easter break centered around the 24th of April in 2011. The data regarding traveling does not specify to which latitude, for how long, or estimate the grade of natural UVB irradiance.

### 2.12 Statistical Analysis

Statistical analyses were performed by using R version 4.1.2 and R Studio built 382.

Homophily, Xi² tables, and bootstrapping with 1000 simulations were used to evaluate the similarities in solarium habits using a simple t-testing for theoretical same-to-same relationships against the simulated same-to-same relationships; as we have done in previous work [65]. Logistic regression was used to compare each person’s 25OHD levels (high/low) and the number of friends with high 25OHD levels. For both the general population, and for each high school individually.

Xi² tables, two-sided Welch’s t-test when two categories are present, and a one-way ANOVA for the rest of the variables, were performed to determine statistically significant differences between groups. Bonferroni correction was applied for high numbers of multiple comparisons. Univariate regression models were used to compare relevant blood serum variables.

## 3 Results

### 3.1 Preliminary analysis of high school biases

There is a heterogeneous distribution of high or low melanin individuals across different high schools (supplementary table 6) with no significant bias (p-value = 0.5). The diet only has a bias in lean fish consumption (p-value = 0.003), with H1 and H8 consuming less than the expected frequency, and H3 and H4 consuming more than expected.

Blood extraction by date (supplementary figure 5) divides the school into Autumn 2010 (H1, H2), Winter 2010 (H3, H7, H8), and Spring 2011 (H4, H5, H6). Schools H2, H7, and H8 also have a significant number of students with blood samples taken during the polar night. H1 and H6 show a bias towards sunbathing traveling (supplementary table 14). H1 was the first school to be tested so it is expected that students had recent travels due to the summer holiday. H6 students are expected to travel more related to training or competitions. The differences in 25OHD levels between solarium and non-solarium users are described further down.

### 3.2 Population levels of vitamin D

Previous studies done within the same population have shown an increase of 25OHD levels associated with vitamin and mineral supplements, physical activity, sunbathing holidays, and use of solariums for both sexes, while only men showed increased levels for the use of snuff, consumption of fortified milk, and fish liver oil [5]. Other studies in the general Tromsø population have shown that vitamin D has been positively associated with older age, blood sample time collection, sunbathing holiday, higher alcohol intake, use of fish oil and vitamin supplements, and negatively associated with smoking and obesity [66]. Adolescents in Tromsø have a higher prevalence of vitamin D deficiency compared with a similar-aged population in Spain [67]. We show 25OHD levels for comparison across categories of all variables of interest (table 1), however, the relevant variables in the general population do not seem to be relevant once we analyze high schools one by one.

**Table 1:**
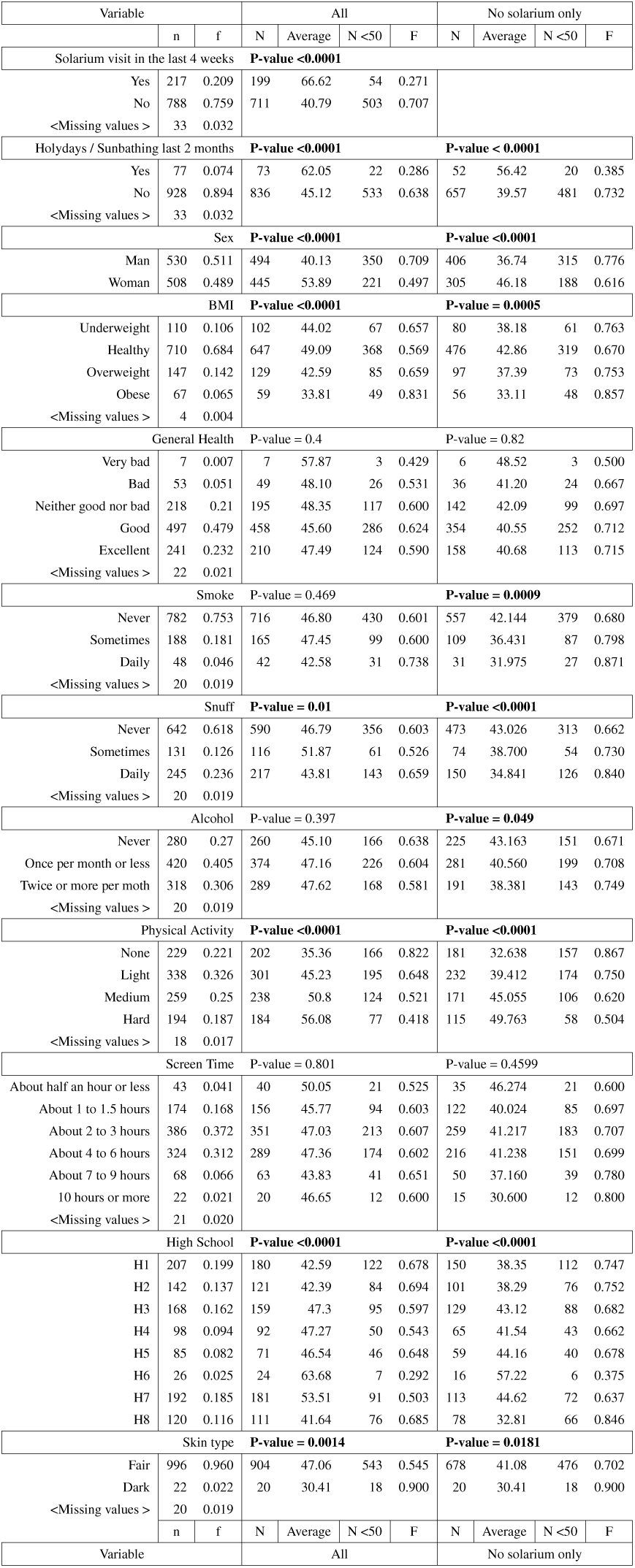
Descriptive statistics with respect to all variables of interest. In the column “Variable” we show the population composition, the column “All” is the analysis of the whole population without missing values, and the column “Non-solarium only” represents the population that did not go to the solarium in the last 4 weeks. In “Variable”, we show the categories of each variable (i.e.: Man, Woman in Sex), the absolute frequency of each category (n), and the relative frequency of each category (f). In both “All” and “Non-solarium only”, we show the absolute frequency of people with valid 25OHD values (N), the average for each category (Average), how many values are below 50 nmol/l (N<50), and the relative frequency in each category of people with low vitamin D (F). To calculate the p-values we run a two-sided Welch’s t-test when two categories are present, and a one-way ANOVA for the rest of the variables. Relevant variables and p-values are highlighted in bold. The Fit Futures 1 study.

| Variable |  |  | All |  |  |  | No solarium only |  |  |  |
| --- | --- | --- | --- | --- | --- | --- | --- | --- | --- | --- |
|  | n | f | N | Average | N<50 | F | N | Average | N<50 | F |
| Solarium visit in the last 4 weeks |  |  | P-value <0.0001 |  |  |  |  |  |  |  |
| Yes | 217 | 0.209 | 199 | 66.62 | 54 | 0.271 |  |  |  |  |
| No | 788 | 0.759 | 711 | 40.79 | 503 | 0.707 |  |  |  |  |
| <Missing values > | 33 | 0.032 |  |  |  |  |  |  |  |  |
| Holidays / Sunbathing last 2 months |  |  | P-value <0.0001 |  |  |  | P-value < 0.0001 |  |  |  |
| Yes | 77 | 0.074 | 73 | 62.05 | 22 | 0.286 | 52 | 56.42 | 20 | 0.385 |
| No | 928 | 0.894 | 836 | 45.12 | 533 | 0.638 | 657 | 39.57 | 481 | 0.732 |
| <Missing values > | 33 | 0.032 |  |  |  |  |  |  |  |  |
| Sex |  |  | P-value <0.0001 |  |  |  | P-value <0.0001 |  |  |  |
| Man | 530 | 0.511 | 494 | 40.13 | 350 | 0.709 | 406 | 36.74 | 315 | 0.776 |
| Woman | 508 | 0.489 | 445 | 53.89 | 221 | 0.497 | 305 | 46.18 | 188 | 0.616 |
| BMI |  |  | P-value <0.0001 |  |  |  | P-value = 0.0005 |  |  |  |
| Underweight | 110 | 0.106 | 102 | 44.02 | 67 | 0.657 | 80 | 38.18 | 61 | 0.763 |
| Healthy | 710 | 0.684 | 647 | 49.09 | 368 | 0.569 | 476 | 42.86 | 319 | 0.670 |
| Overweight | 147 | 0.142 | 129 | 42.59 | 85 | 0.659 | 97 | 37.39 | 73 | 0.753 |
| Obese | 67 | 0.065 | 59 | 33.81 | 49 | 0.831 | 56 | 33.11 | 48 | 0.857 |
| <Missing values > | 4 | 0.004 |  |  |  |  |  |  |  |  |
| General Health |  |  | P-value = 0.4 |  |  |  | P-value = 0.82 |  |  |  |
| Very bad | 7 | 0.007 | 7 | 57.87 | 3 | 0.429 | 6 | 48.52 | 3 | 0.500 |
| Bad | 53 | 0.051 | 49 | 48.10 | 26 | 0.531 | 36 | 41.20 | 24 | 0.667 |
| Neither good nor bad | 218 | 0.21 | 195 | 48.35 | 117 | 0.600 | 142 | 42.09 | 99 | 0.697 |
| Good | 497 | 0.479 | 458 | 45.60 | 286 | 0.624 | 354 | 40.55 | 252 | 0.712 |
| Excellent | 241 | 0.232 | 210 | 47.49 | 124 | 0.590 | 158 | 40.68 | 113 | 0.715 |
| <Missing values > | 22 | 0.021 |  |  |  |  |  |  |  |  |
| Smoke |  |  | P-value = 0.469 |  |  |  | P-value = 0.0009 |  |  |  |
| Never | 782 | 0.753 | 716 | 46.80 | 430 | 0.601 | 557 | 42.144 | 379 | 0.680 |
| Sometimes | 188 | 0.181 | 165 | 47.45 | 99 | 0.600 | 109 | 36.431 | 87 | 0.798 |
| Daily | 48 | 0.046 | 42 | 42.58 | 31 | 0.738 | 31 | 31.975 | 27 | 0.871 |
| <Missing values > | 20 | 0.019 |  |  |  |  |  |  |  |  |
| Snuff |  |  | P-value = 0.01 |  |  |  | P-value <0.0001 |  |  |  |
| Never | 642 | 0.618 | 590 | 46.79 | 356 | 0.603 | 473 | 43.026 | 313 | 0.662 |
| Sometimes | 131 | 0.126 | 116 | 51.87 | 61 | 0.526 | 74 | 38.700 | 54 | 0.730 |
| Daily | 245 | 0.236 | 217 | 43.81 | 143 | 0.659 | 150 | 34.841 | 126 | 0.840 |
| <Missing values > | 20 | 0.019 |  |  |  |  |  |  |  |  |
| Alcohol |  |  | P-value = 0.397 |  |  |  | P-value = 0.049 |  |  |  |
| Never | 280 | 0.27 | 260 | 45.10 | 166 | 0.638 | 225 | 43.163 | 151 | 0.671 |
| Once per month or less | 420 | 0.405 | 374 | 47.16 | 226 | 0.604 | 281 | 40.560 | 199 | 0.708 |
| Twice or more per moth | 318 | 0.306 | 289 | 47.62 | 168 | 0.581 | 191 | 38.381 | 143 | 0.749 |
| <Missing values > | 20 | 0.019 |  |  |  |  |  |  |  |  |
| Physical Activity |  |  | P-value <0.0001 |  |  |  | P-value <0.0001 |  |  |  |
| None | 229 | 0.221 | 202 | 35.36 | 166 | 0.822 | 181 | 32.638 | 157 | 0.867 |
| Light | 338 | 0.326 | 301 | 45.23 | 195 | 0.648 | 232 | 39.412 | 174 | 0.750 |
| Medium | 259 | 0.25 | 238 | 50.8 | 124 | 0.521 | 171 | 45.055 | 106 | 0.620 |
| Hard | 194 | 0.187 | 184 | 56.08 | 77 | 0.418 | 115 | 49.763 | 58 | 0.504 |
| <Missing values > | 18 | 0.017 |  |  |  |  |  |  |  |  |
| Screen Time |  |  | P-value = 0.801 |  |  |  | P-value = 0.4599 |  |  |  |
| About half an hour or less | 43 | 0.041 | 40 | 50.05 | 21 | 0.525 | 35 | 46.274 | 21 | 0.600 |
| About 1 to 1.5 hours | 174 | 0.168 | 156 | 45.77 | 94 | 0.603 | 122 | 40.024 | 85 | 0.697 |
| About 2 to 3 hours | 386 | 0.372 | 351 | 47.03 | 213 | 0.607 | 259 | 41.217 | 183 | 0.707 |
| About 4 to 6 hours | 324 | 0.312 | 289 | 47.36 | 174 | 0.602 | 216 | 41.238 | 151 | 0.699 |
| About 7 to 9 hours | 68 | 0.066 | 63 | 43.83 | 41 | 0.651 | 50 | 37.160 | 39 | 0.780 |
| 10 hours or more | 22 | 0.021 | 20 | 46.65 | 12 | 0.600 | 15 | 30.600 | 12 | 0.800 |
| <Missing values > | 21 | 0.020 |  |  |  |  |  |  |  |  |
| High School |  |  | P-value <0.0001 |  |  |  | P-value <0.0001 |  |  |  |
| H1 | 207 | 0.199 | 180 | 42.59 | 122 | 0.678 | 150 | 38.35 | 112 | 0.747 |
| H2 | 142 | 0.137 | 121 | 42.39 | 84 | 0.694 | 101 | 38.29 | 76 | 0.752 |
| H3 | 168 | 0.162 | 159 | 47.3 | 95 | 0.597 | 129 | 43.12 | 88 | 0.682 |
| H4 | 98 | 0.094 | 92 | 47.27 | 50 | 0.543 | 65 | 41.54 | 43 | 0.662 |
| H5 | 85 | 0.082 | 71 | 46.54 | 46 | 0.648 | 59 | 44.16 | 40 | 0.678 |
| H6 | 26 | 0.025 | 24 | 63.68 | 7 | 0.292 | 16 | 57.22 | 6 | 0.375 |
| H7 | 192 | 0.185 | 181 | 53.51 | 91 | 0.503 | 113 | 44.62 | 72 | 0.637 |
| H8 | 120 | 0.116 | 111 | 41.64 | 76 | 0.685 | 78 | 32.81 | 66 | 0.846 |
| Skin type |  |  | P-value = 0.0014 |  |  |  | P-value = 0.0181 |  |  |  |
| Fair | 996 | 0.960 | 904 | 47.06 | 543 | 0.545 | 678 | 41.08 | 476 | 0.702 |
| Dark | 22 | 0.022 | 20 | 30.41 | 18 | 0.900 | 20 | 30.41 | 18 | 0.900 |
| <Missing values > | 20 | 0.019 |  |  |  |  |  |  |  |  |
|  | n | f | N | Average | N<50 | F | N | Average | N<50 | F |
| Variable |  |  | All |  |  |  | No solarium only |  |  |  |

“Solarium visit in the last 4 weeks” shows a dramatic difference in vitamin D deficiency between people who visit solariums (27.1%) and people who do not (70.7%). As such, all further analysis regarding high school bias is performed using only people who do not go to the solarium (supplementary tables 8, 9, 10, 11, 12 and 13).

“BMI” is known to be a risk factor for vitamin D levels. Fat cells can sequester vitamin D due to being a fat-soluble vitamin, and it is not surprising to find lower levels for higher %fat individuals. This population has a similar BMI for both men (22.51 ± 4.22) and women (22.62 ± 4.24) in the total population and for men (22.61 ± 4.38) and women (22.89 ± 4.48) in the non-solarium population. However, BMI is not evenly distributed across high schools for non-solarium users (p-value < 0.0001). There is a higher prevalence of obesity in H2 and H5, overweight in H8, and underweight in H4. H7 has a lower prevalence of overweight and obese individuals. All H6 students fall into the “Healthy” BMI category. H6 total number of students is low, and the p-value to ascertain a non-random habit is not significant. However, H6 is exclusively a sports high-school so it is likely that they are biased towards a healthier lifestyle even though the binomial test for this variable cannot suggest so.

“Sex” differences are partially explained because women visit the solarium more often than men and this influence will be discussed later. High schools are biased with respect to the sex distribution (p-value < 0.0001), with H1 and H8 leaning toward men, and H2 and H3 leaning toward women.

Recreational drug consumption also shows a strong bias towards high school for alcohol, smoking, and snuff habits (p-value < 0.0001 for all cases). Schools that show a pro-smoking bias are H1, H2, H5, and H8. Schools that show a pro-snuff bias are in H1 and have very strong usage in H8. Schools that show a pro-alcohol bias are H5 and very strong in H8. Anti-alcohol bias is shown in H6. Anti-snuff bias is shown in H7, and very slightly in H3 and H6. There seems to be no “no smoking” bias, but smoke frequency is lower in H3, H4, and H7. In H6 nobody smokes, but once again the p-value is not low enough to suggest a non-random habit, although H6 is a sport school so it is likely that they avoid smoking.

Physical activity also shows a bias with respect to high schools (p-value < 0.0001). With H2 and H8 leaning toward none, H1, H3, and H5 toward light to medium, and H6 and H7 toward hard.

Finally, we stratify all non-solarium users by high schools and run a two-sided Welch’s t-test when two categories are present, and a one-way ANOVA for the rest of the variables. After correcting all results for multiple testing with Bonferroni, “Sex” was relevant for H8 (p-value = 0.0059, {men n = 59, x = 28.45 nmol/l; women n = 19, x = 46.34 nmol/l}), “Physical Activity” for H3 (p-value = 0.03, {none = 24, x = 30.89 nmol/l; light n = 44, x = 40.14 nmol/l; medium n = 42, x = 49.47 nmol/l; hard n = 19, x = 51.2 nmol/l}). *“Holiday / Sunbathing in the last 2 months”* was only significant in H1, first school to be tested after summer, and H3, first school to be completely tested after Christmas.

### 3.3 Similarities in solarium habits among friends

Women in the general population have higher vitamin D levels across the year (table 1 and figure 1), this also happens with people going to the solarium (table 1 and figure 2). We checked Xi² tables for diet and solarium habits to test differences between men and women. The diet table showed no significant differences in diet, but significant differences in solarium habits were found (p-value < 0.0001) (table 2). Men tend to not go to the solarium (33.64% of the solarium population) while women tend to go to the solarium (66.46%). This might indicate that social relationships between men and women groups affect solarium habits.

**Figure 1:**
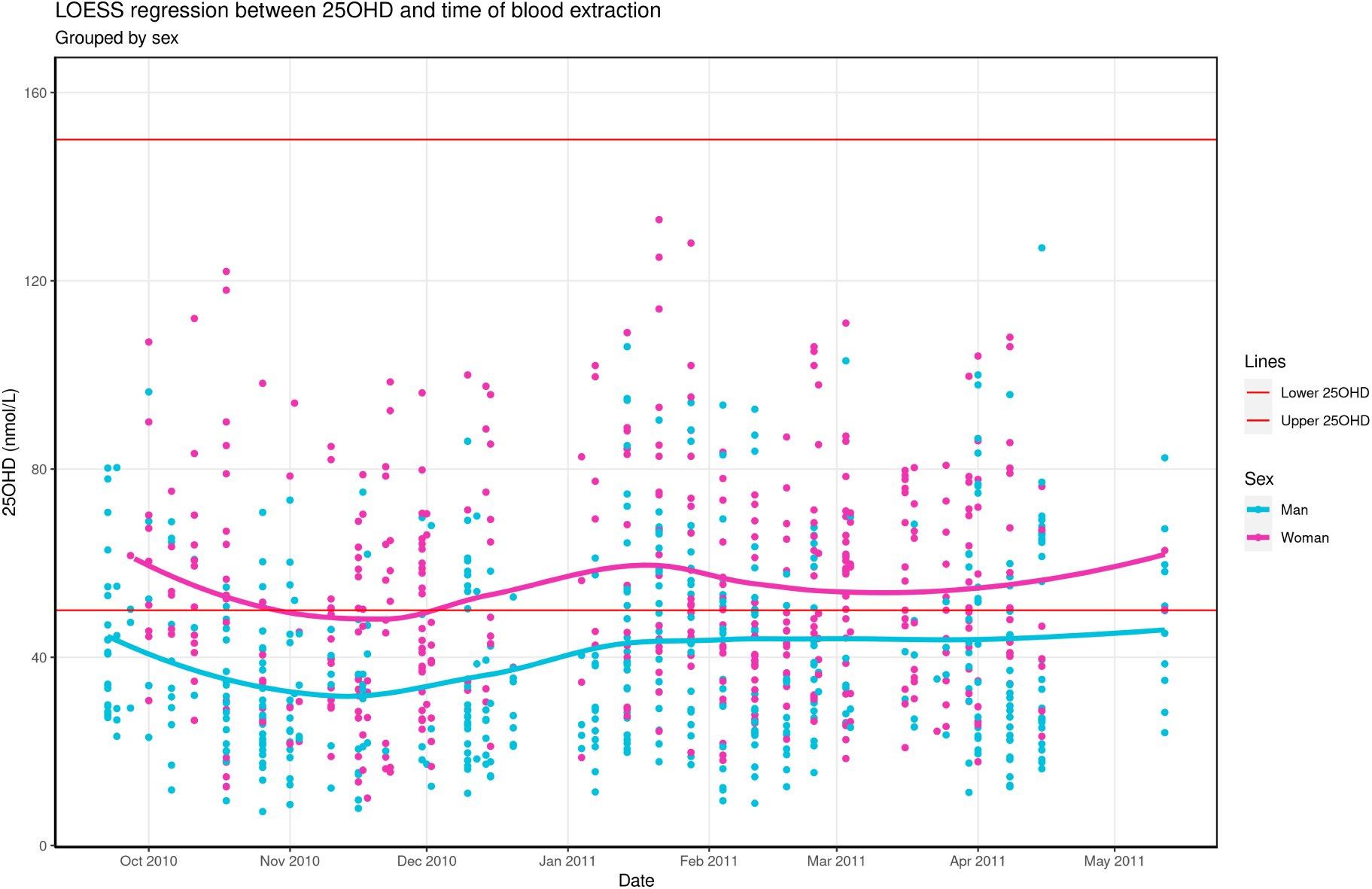
25OHD levels from October 2010 to May 2011 divided by sex. Horizontal red lines mark the boundaries of healthy 25OHD levels. Women display higher levels than men across the whole year. The Fit Futures 1 study, N = 890.

**Figure 2:**
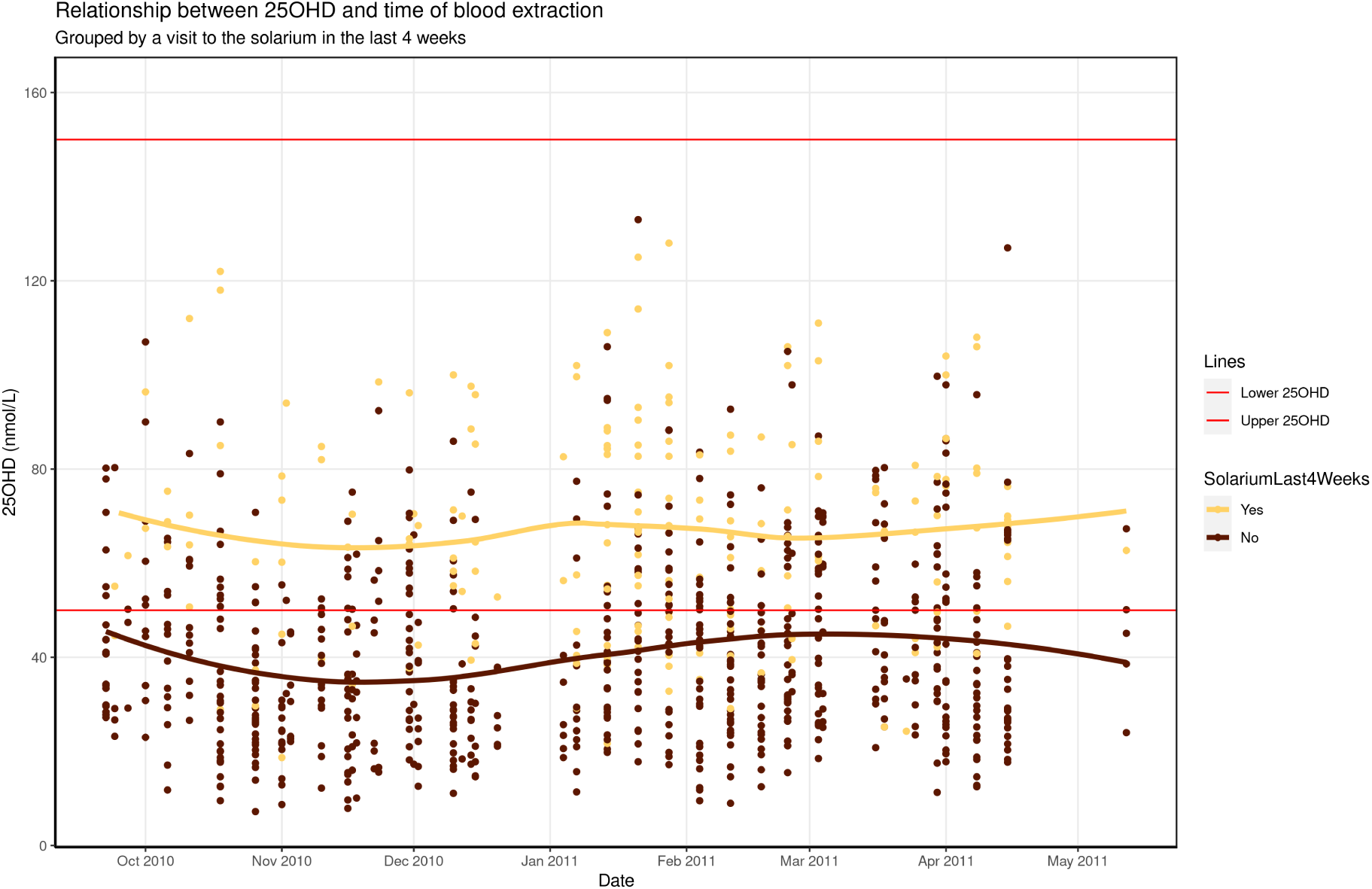
25OHD levels from October 2010 to May 2011 divided by solarium habit. Horizontal red lines mark the boundaries of healthy 25OHD levels. People going to the solarium display higher levels than people who don’t go to the solarium across the whole year. People who don’t go to the solarium display a dip in levels in December while a peak of levels in near April. There was a second dip in the middle of May due to a small sample size and low vitamin D levels of students who were tested at that time. The Fit Futures 1 study, N = 890

**Table 2:** Xi² table for “Did you go to the solarium in the last 4 weeks?” (yes/no), and sex. The header numbers indicate the total population absolute and relative frequency for men and women. The lower marginals are how many of these have available data with respect to the solarium question. Each inner cell is divided into 3 parts; the left-most is the total number of relationships in this combination, the center one is the expected number of relationships, and the right part contains an arrow indicating over (up) or underrepresented (down) using a two-sided binomial test with at least p-value < 0.1. Women are biased toward the yes answer, while men are biased towards the no answer. The Fit Futures 1 study.

| Xi² <0.0001 | Man |  |  | Woman |  |  | Total | Freq |
| --- | --- | --- | --- | --- | --- | --- | --- | --- |
|  | n = 530 , f = .51 |  |  | n = 508 , f = .49 |  |  |  |  |
| Yes | 73 | (106) | ↓ | 144 | (103) | ↑ | 217 | 21% |
| No | 436 | (386) | ↑ | 352 | (376) | ↓ | 788 | 76% |
| Total | 509 |  |  | 496 |  |  | 1005 |  |
| Frequency | 49% |  |  | 48% |  |  | 97% |  |

Sex is a strong homophily variable defining friendship (84.05%), and men are friends with mostly men (72.06%) and women are friends with mostly women (72.89%). Our previous studies [65] on the data also show that men have fewer friends (3.37) on average than women (3.85) (p-value = 0.02). The homophily for solarium visitors is 68.36%, with “yes” having a homophily of 22% but not significant, and “no” having a 66% (8% lower than it should) with p-value < 0.0001. This means that people who do not go to the solarium are less likely to form friendships with each other. Moreover, it correlates with the sex dynamics of men having fewer friends, and men not going into the solarium as often.

We compared the social network against simulated networks to check if people going to the solarium influence other people into going to the solarium. For this analysis, we filtered out people who did not answer the question about the solarium (n = 33). Instead of the original 3767 relationships, we were left with 3575 relationships in total. Doing 1000 simulations we got an average of 2365 same-to-same relationships with a significance of p-value = 0.008 for the overall network. This indicates that people are biased and relationships with respect to solarium habits are non-random. We tried the same analysis again using only same-sex friends. Sex has very high homophily, as such we believe that barely any influence from men to women and vice-versa is lost in this analysis. For women, the total same-to-same relationships are 1049 with a p-value of 0.0002 which seems to indicate that women tend to form groups of friends with the same solarium habits. For men, however, we have 1122 same-to-same relationships, resulting in a p-value of 0.23, showing no bias in this case.

### 3.4 Social influence in 25OHD

We want to check if non-solarium students with high 25OHD levels influence other non-solarium students to have high 25OHD levels as well by using logistic regression. The result shows that the chances of having high vitamin D increase by 7.25% [5.65%, 8.85%] for each additional friend who also has high vitamin D (figure 3). This result however can be biased. For example, different high schools had different blood extraction dates around the year, and friendship has very high homophily for high school (87.5%), so it is possible that we are just measuring high schools with higher density connections, while those schools coincidentally extracted blood at peak or bottom sun irradiance.

**Figure 3:**
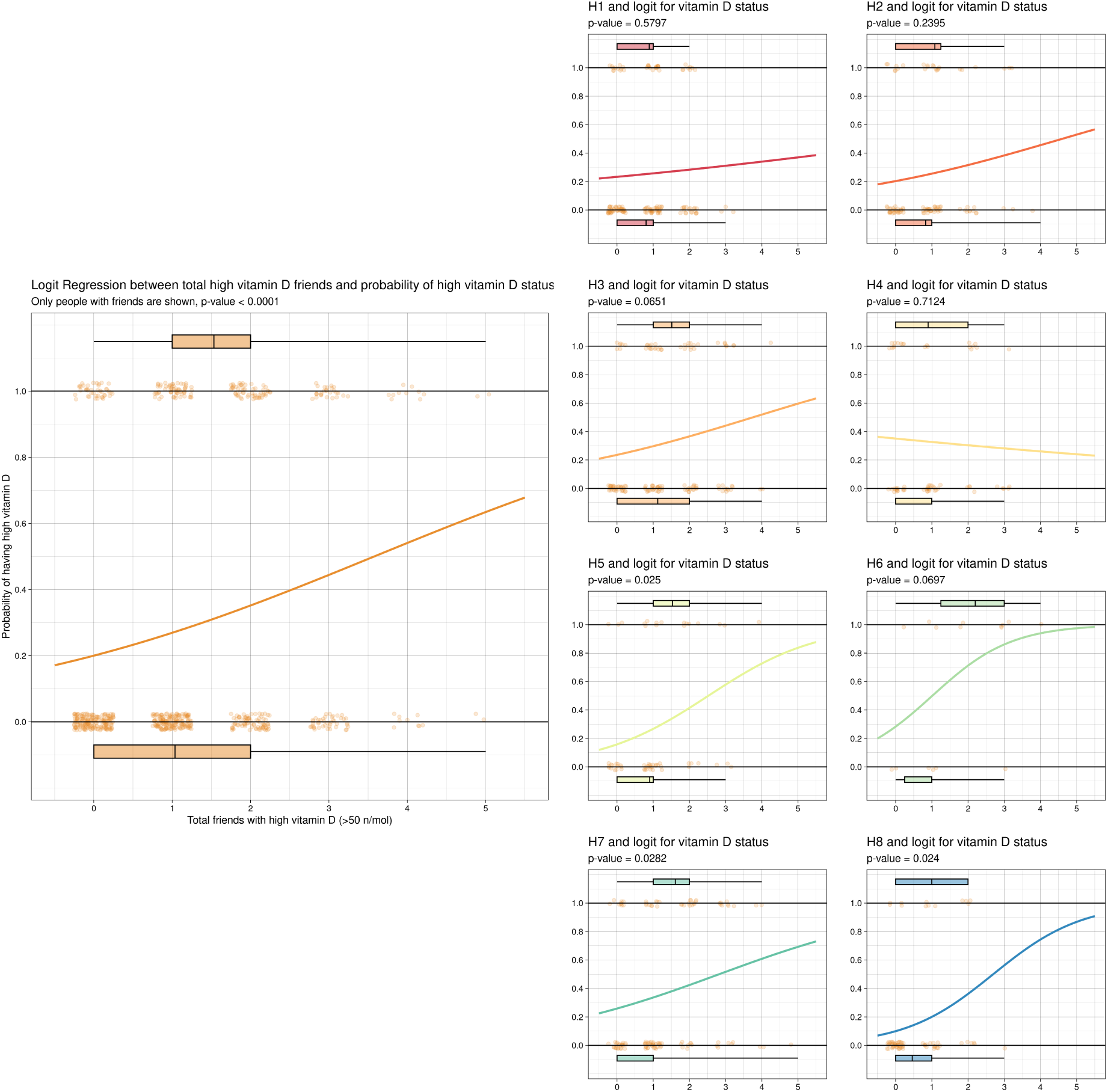
(Left) Logistic regression with respect to 25OHD levels between each person and high 25OHD friends. Each dot represents a person who does not go to the solarium, and who has at least one friend. The dots are laid out on the Y-axis in 1 if the subject has high vitamin D (25OHD > 50 nmol/l), or 0 if low vitamin D (25OHD < 50 nmol/l). In the X-axis we count how many high vitamin D friends this person has. The boxplots represent the difference in the number of high vitamin D friends between people who have low vitamin D (0) or high vitamin D (1). People with low vitamin D have an average of 1.04 friends with high vitamin D, while people with high vitamin D have an average of 1.53 friends with high vitamin D (p-value < 0.0001). (Right) The logistic regression analysis was performed for each high school independently. Each plot has a p-value displayed under the title. Relevant levels are for H3, H5, H6, H7 and H8.

We repeated the logistic regression analysis for each high school independently. Five of the eight also showed statistically significant 25OHD differences (figure 3) (H3, H5, H6, H7, and H8), with two having blood extraction during the polar night (H7 and H8). This suggests that friends influence each other into sharing certain lifestyles which makes 25OHD levels similar.

Furthermore, we plotted each student’s 25OHD level against his or her friends’ average of 25OHD. We see a weak correlation (R²= 0.1, p-value < 0.0001) among the whole population (supplementary figure 17). The same analysis of each individual high school shows more relevant results for H2, H5, and H8 (supplementary figure 17). We also performed another 1000 simulations for each high school individually, for non-solarium users only, and found non-random friendship based on vitamin D status for H1, H2, H3, H4, H5, and H8 (supplementary table 5).

### 3.5 Levels by nutritional data

Finally, we wanted to check if the diet was influenced by friends. Performing ANOVA in all our nutritional variables showed no 25OHD significance difference between consumption groups for fat fish, cheese, dairy, vitamin pills, or even fish oil complements. Analyzing each high school independently, we found no correlation at all in any of the 48 possible combinations without even applying any post-hoc correction. The only difference present was in the consumption of lean fish in the general population, however, the difference was only between lower-frequency consumption groups and the middle-frequency consumption groups, while the high-frequency groups show approximately the same levels of 25OHD as people not eating any lean fish. We believe that this is the effect of confounding variables between lean fish frequency and 25OHD. We also tried to perform the same 1000 simulations for each of the 6 food groups in the overall network and we found no evidence that friends influence dietary habits among each other.

## 4 Discussion

### 4.1 Social influence of high school in vitamin D levels

Among the variables of sex, BMI, recreational drugs, health, solarium, and sports habits, high school has the stronger homophily when it comes to friendship (min 70% per school). We see that women tend to form friendships with other women if they share the same solarium habits. In previous works [58] the main motivation of Norwegian teenagers for going to the solarium was *“To get a tan”* with nearly 80% for girls and 60% for boys in both 2016 and 2017, followed by *“To prepare for holidays”* 32% in girls and 16.3% in boys in 2016, and near 23% for both in 2017. *“To make vitamin D”* appears as the third motivation with 20% for both in 2016 and 17.5% for both in 2017. It should be noticed that previous work showed that none of these reasons are justified due to solariums being too powerful, having the wrong UVA/UVB ratio, not increasing protection, and increasing hypervitaminosis D [55, 58, 68–70].

For non-solarium users, it appears that there is a general tendency in the population in which students who have a higher number of friends with high vitamin D levels tend to have higher vitamin D levels themselves. This tendency is also present in 5 of the 8 high schools. The simulations and the regression models also show a tendency to have similar vitamin D levels compared to friends. This accounts for possible bias towards total UVB. Furthermore, skin tone seems to be homogenously distributed across high schools, and the total number of dark-skin individuals is low (2% population) so this effect is not due to having an unbalanced skin type distribution. In our data, there seems to be no bias towards diet in any school other than lean fish consumption. Lean fish does not contain significant levels of vitamin D, and higher 25OHD levels in this consumption group appear to be due to confounding factors.

This seems to indicate that people follow the same healthy or unhealthy habits related to vitamin D absorption as their friends. This might include socio-economic factors such as being able to afford more traveling to more sunny areas, being more physically active and spending more time outdoors, having better diets that are rich in fatty fish, or better education related to vitamin D deficiency and supplementation.

### 4.2 Confounding of high school in vitamin D levels with other factors

H8 is the school with the lowest average 25OHD levels and is biased towards all the variables with lower-than-average 25OHD levels (males with −9.44 nmol/l 25OHD lower than women, not eating lean fish −5.71, alcohol consumption −3.49, smoking −6.7, snuff −6.91, and no physical activity −10.93). H1 is a similar school with no bias towards alcohol, and bias towards light physical activity; that despite having more traveling than any other school, is tied at second place with lower 25OHD levels. These two schools have 31% of the student population. In contrast, H6 and H7 are the complete opposite, with the highest vitamin D levels of any school, and bias towards no alcohol, no smoke, no snuff, and hard physical activity. Both H6 and H7 alone represent 21% of the population. Studying these variable levels for each high school independently, among students that do not go to the solarium, shows only 4 significant values after Bonferroni correction. H8 for sex, H3 for physical activity, and H1 and H3 for traveling.

Non-solarium women in H8 have +13.77 nmol/l average vitamin D levels than their male counterparts. These women do not show different habits than men in diet, BMI, physical activity, or recent sunbathing. This school has a bias towards the male student population, but neither sex displays different social dynamics that differ significantly from any other school. A network plot from the non-solarium population in H8 (supplementary figure 16), confirms the logistic regression analysis and shows a pattern in which women with high vitamin D are friends with other women with high vitamin D, low vitamin D men are friends with low vitamin D friends, and high vitamin D men are friends with high vitamin D friends. Therefore, a proper follow-up of the students in this school can help to understand what helped these women to gain much better levels than their male counterparts, which is of particular interest given that H8 blood samples were taken in the middle of the polar night. A possible explanation is the use of oral contraceptives with estrogen [71], which are common in this school. However, in this school, we found no 25OHD significant difference between women not going to the solarium using Microgynon (n = 8) and those not using oral contraceptives (n = 9). The rest of the network plots, for all other high schools, also show clusters of non-solarium men or women with similar 25OHD levels in accordance to the simulation results (supplementary figures 9, 10, 11, 12, 13, 14, 15)

Physical activity (PA) in H3 is the only place where this variable is significant. PA is linked to outdoor sports and activities. However, Tromsø is cold and has a daily average temperature ranging from an average of 7.8°C in September to –3.3°C in February. Due to both clothing and lack of UVB, it is very unlikely that people in this school are exposed to enough skin to get enough UVB on a daily basis. However, it is likely that people doing physical activity have a healthier diet. Evidence suggests that obese people require higher vitamin D [72, 73], so physical activity PA is recommended to lower the vitamin D requirements regardless of whether it helps absorption or not.

Traveling to southern latitudes also helps with the UVB levels in H1 and H3. However, we do not have refined data regarding the amount of time of sun exposure, latitude, or possible skin lesions during that time to make a proper assessment. Further data is needed to evaluate the risk-benefit relationship of sun exposure with respect to gains of vitamin D in this particular population. Food fortification seems to have helped with lowering cancer mortality rates in Europe [59], and will be a safer public health approach than recommending increasing UVA+UVB exposure.

Altogether the results from the present analysis indicate that the only good predictor of vitamin D levels in this population is UVB radiation absorption due to associations with the date of the year, traveling, or solarium usage, and the significance of vitamin D with respect to these variables stems from the biases present in high school social dynamics.

### 4.3 Contradictory results and studies

We showed that getting closer to people with high vitamin D levels tends to increase vitamin D levels in the individual. This is of course due to similar interests to those lifestyles of peers, and we have shown extensively the effects it has in different high schools. However, due to the limited nutritional data and PA estimates, we cannot ascertain the effects that each possible healthier lifestyle may have on the network.

Vitamin D research presents contradictory evidence in many studies. This might be due to 25OHD tests not being properly standardized and poor experiment design [7, 74, 75] or not accounting for other supplements [76, 77]. Our study also adds to the confusion regarding the effective sources of vitamin D. For dietary habits in each school independently, or in the total population, there are no significant differences in vitamin D levels with neither fat fish consumption, and for daily intake of fish oil pills that typically contain 500-1.000 UI of D3 per pill. It is very unlikely that people reporting taking these pills do not display higher levels. For example, an interventional study in Northern Ireland with university students showed that daily supplements of 600 IU increased by almost +40nmol/l with respect to the control group [78]. Our dataset uses memory-based dietary assessment methods (M-BMs), described as *“uncritical faith in the validity and value of M-BMs has wasted substantial resources and constitutes the greatest impediment to scientific progress in obesity and nutrition research”* [79]. Similar conclusions can be found in self-reported data in the geriatric residential population [80]. Other studies suggest that the validity of Automated Self-Administered 24-hour recalls (ASA24s) is good enough [81] and multiple ASA24s and 4-d food records (4DFRs) provided the best estimates [82]. Our students report on food-frequency questionnaires (FFQs), which vary from the last 24 hours of food consumption prior to blood extraction, which was not even taken after 8 hours of fasting. It is important to enhance the scrutiny of dietary intake in future epidemiological studies. As such, we believe using FFQ is not a good option, and at the very least, having a nutritional professional oversee the ASA24s. Ideally, 4DFRs should be inputted into a food database (such as https://www.matportalen.no/) to retrieve final nutritional values and compare them with biomarkers and metabolites in blood.

Similarly, PA can be collected in a better way. The metabolic equivalent of task (MET) is a unit that measures how active a person is and roughly translates into 1 kcal/kg/hour, 1W/kg, or the energy required to sit down for an hour. In our questionnaires, PA is also self-reported and open to interpretation. Light activity includes walking and cycling, which vary from 2 to 6 METs. Medium activity includes recreational sports, such as light weightlifting (3.5 METs), tennis (5), basketball (8), football (10), jogging (11), or snow cleaning (4-8). Hard includes sports competitions, which again, vary too much for each type. Our categories have potential MET overlapping. METs have some limitations, and it is harder to calculate the exact value from person to person, but while it is also self-reported, it is a more objective measure of PA. This would also keep specific activities separated for better analysis. Alternatively, we could make use of accelerometers as has been reported in previous FF studies [83], but didn’t have access to this data.

## 5 Conclusions

We saw that UVB radiation presents itself as the main influence of 25OHD levels in this population, even though low levels are present due to the Arctic geolocation. Among people not using solarium, those with greater numbers of friends with >50nmol/l tend to have higher levels of 25OHD themselves and vice versa, with an estimated probability of +7.25% per high vitamin D friend. FFQ seems to be a non-reliable tool for nutrition assessment and at the very least needs to be substituted with an ASA24s questionnaire; same with PA and METs.

## 6 List of abbreviations

| Acronym | Meaning |
| --- | --- |
| 25OHD | 25-hydroxycholecalciferol (Calcifediol) |
| 4DFRs | 4-d food records |
| ASA24s | Automated Self-Administered 24-h recalls |
| BMI | Body Mass Index |
| FF | Fit Futures |
| FFQs | food-frequency questionnaires |
| LC-MS/MS | High-pressure liquid chromatography-mass spectroscopy |
| M-BMs | memory-based dietary assessment methods |
| MET | metabolic equivalent of task |
| PA | Physical Activity |
| PTH | Parathyroid hormone |
| REK | Regional Committee of Medical and Health Research Ethics |
| UNN | University Hospital of North Norway |
| UVA | Ultraviolet A |
| UVB | Ultraviolet B |
| VDSP | Vitamin D Standardization Program |

## 7 Declarations

### 7.1 Ethics approval and consent to participate

A declaration of consent was signed by each participant in FF1, participants younger than 16 years of age had to bring written consent from a parent or guardian. FF1 was approved by The Regional Committee of Medical and Health Research Ethics (REK) and the Norwegian Data Protection Authority. The present study was approved by REK North, reference 2011/1710 /REK Nord.

### 7.2 Consent for publication

Not applicable

### 7.3 Availability of data and materials

The Fit Futures data is not publicly available due to Norwegian privacy laws, but researchers can apply for access at https://uit.no/research/fitfutures

The analysis code is open source using the AGPLv3 license and it is available at our GIT repository (https://github.com/uit-hdl/mimisbrunnr/)

Our GIT repository also contains all results displayed here. Formats include Latex, CSV, and HTML for all tables, and PNG and PDF for all images. To avoid visual cluttering, some p-values displayed here are in GP Prism 5.04/d format (asterisks instead of numbers). The raw results contain all tables with both GP Prism format and the numerical format.

### 7.6 Authors contributions

RANC performed conceptualization, methodology, software, formal analysis, data curation, visualization, original draft, review, and editing of the paper. ASF and LAB performed review and editing, supervision, project administration, and funding acquisition. CSN participated in the conceptualization, review and editing, funding acquisition, and project administration. AMH collaborated with review editing and supervision.

## Data Availability

All data produced in the present study are available upon reasonable request to the authors

https://uit.no/research/fitfutures?p_document_id=668286&Baseurl=/research/

## Acknowledgements

We are grateful for the contribution of the participants in the Fit Futures 1 study. We also thank the Clinical Research Department at the University Hospital of North Norway, Tromsø, for data collection. We also want to thank the Department of Microbiology and Infection Control and the Department of Community Medicine at the Faculty of Health Sciences for their analysis of the different blood samples.

We would also like to thank the help and collaboration of Guri Grimmes of the Tromso Endocrine Research Group, Department of Clinical Medicine at the UiT The Arctic University of Norway, and Division of Internal Medicine, University Hospital of North Norway, Tromso, Norway.

## 8 Bibliography

## 9 Supplementary materials

## 10 High-schools information

### 10.1 Map overview

**Figure 4:**
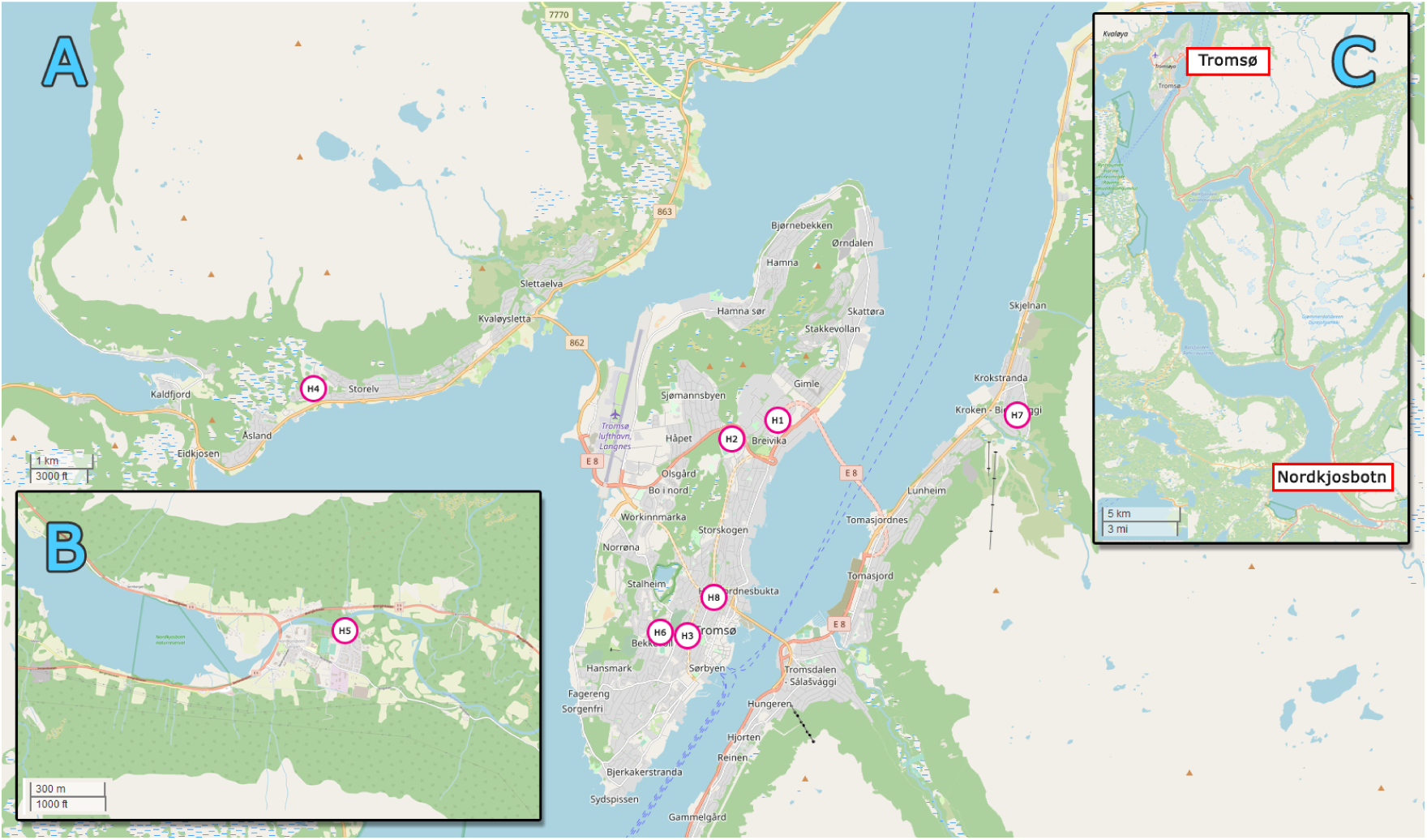
Overview map with the different high schools which were studied. Each high school location is highlighted with a circle with the ID of the school in the center. Area “A” is the view of the Tromsø area, schools H1 to H4 and H6 to H8 are located here. Area “B” is the view of Nordkjosbotn where H5 is located. Area “C” shows the distance and scale between A and B.

### 10.2 High-schools summary table

**Table 3:**
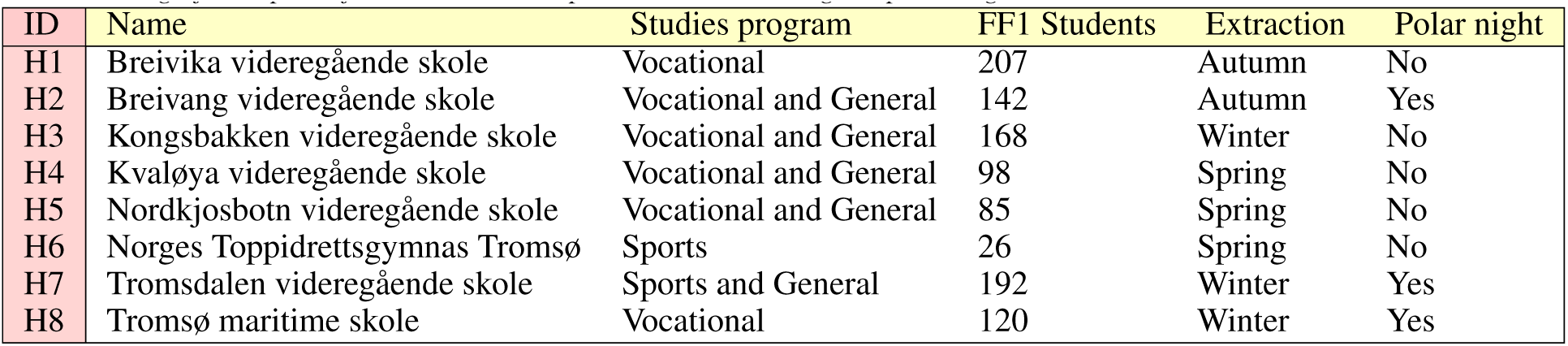
Information with the school names, study program, the total number of students in FF1, astronomical season, and whether a significant part of the student’s samples was taken during the polar night.

| ID | Name | Studies program | FF1 Students | Extraction | Polar night |
| --- | --- | --- | --- | --- | --- |
| H1 | Breivika videregående skole | Vocational | 207 | Autumn | No |
| H2 | Breivang videregående skole | Vocational and General | 142 | Autumn | Yes |
| H3 | Kongsbakken videregående skole | Vocational and General | 168 | Winter | No |
| H4 | Kvaløya videregående skole | Vocational and General | 98 | Spring | No |
| H5 | Nordkjosbotn videregående skole | Vocational and General | 85 | Spring | No |
| H6 | Norges Toppidrettsgymnas Tromsø | Sports | 26 | Spring | No |
| H7 | Tromsdalen videregående skole | Sports and General | 192 | Winter | Yes |
| H8 | Tromsø maritime skole | Vocational | 120 | Winter | Yes |

### 10.3 High-schools dates of blood extraction

**Figure 5:**
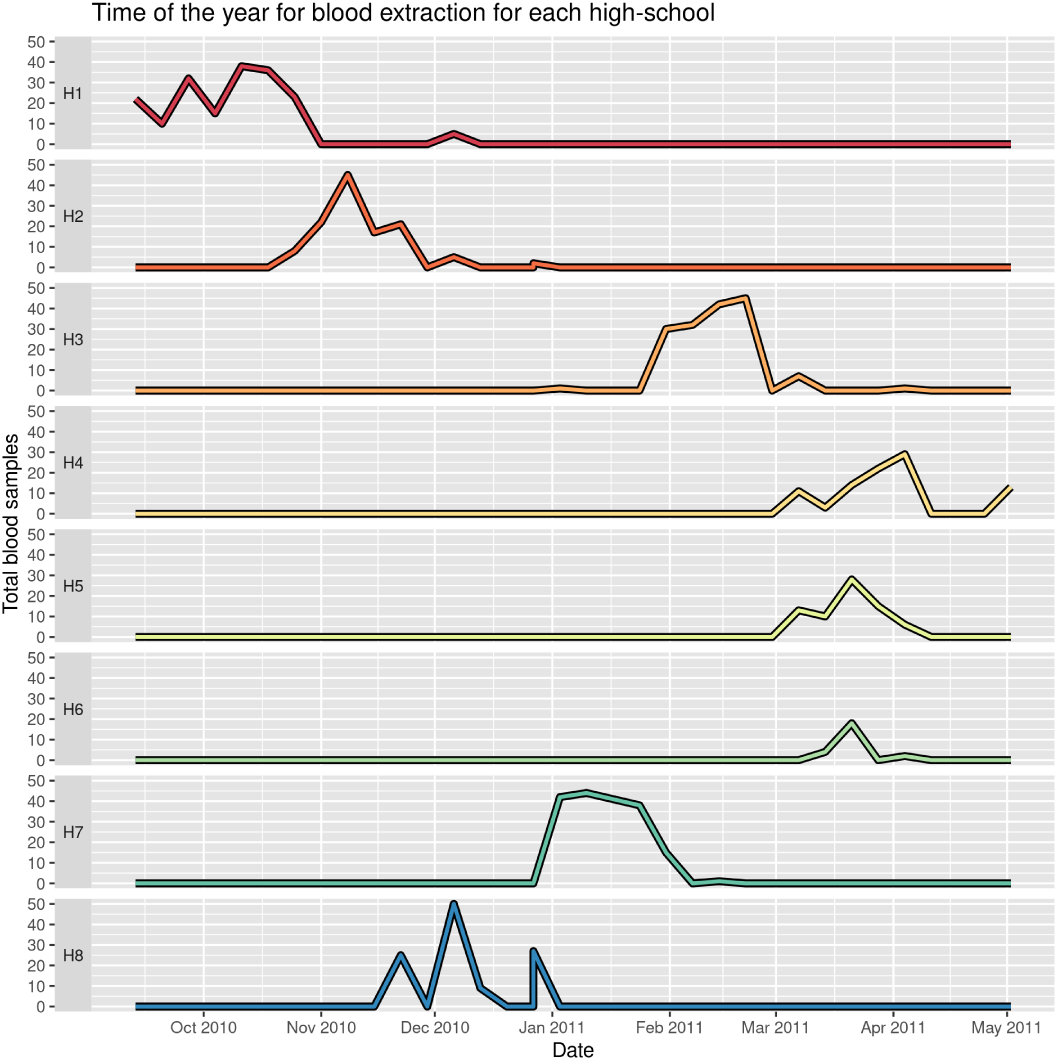
Total blood extractions per high school across time of the year for each high school.

## 11 Networks information

### 11.1 All graphs

**Figure 6:**
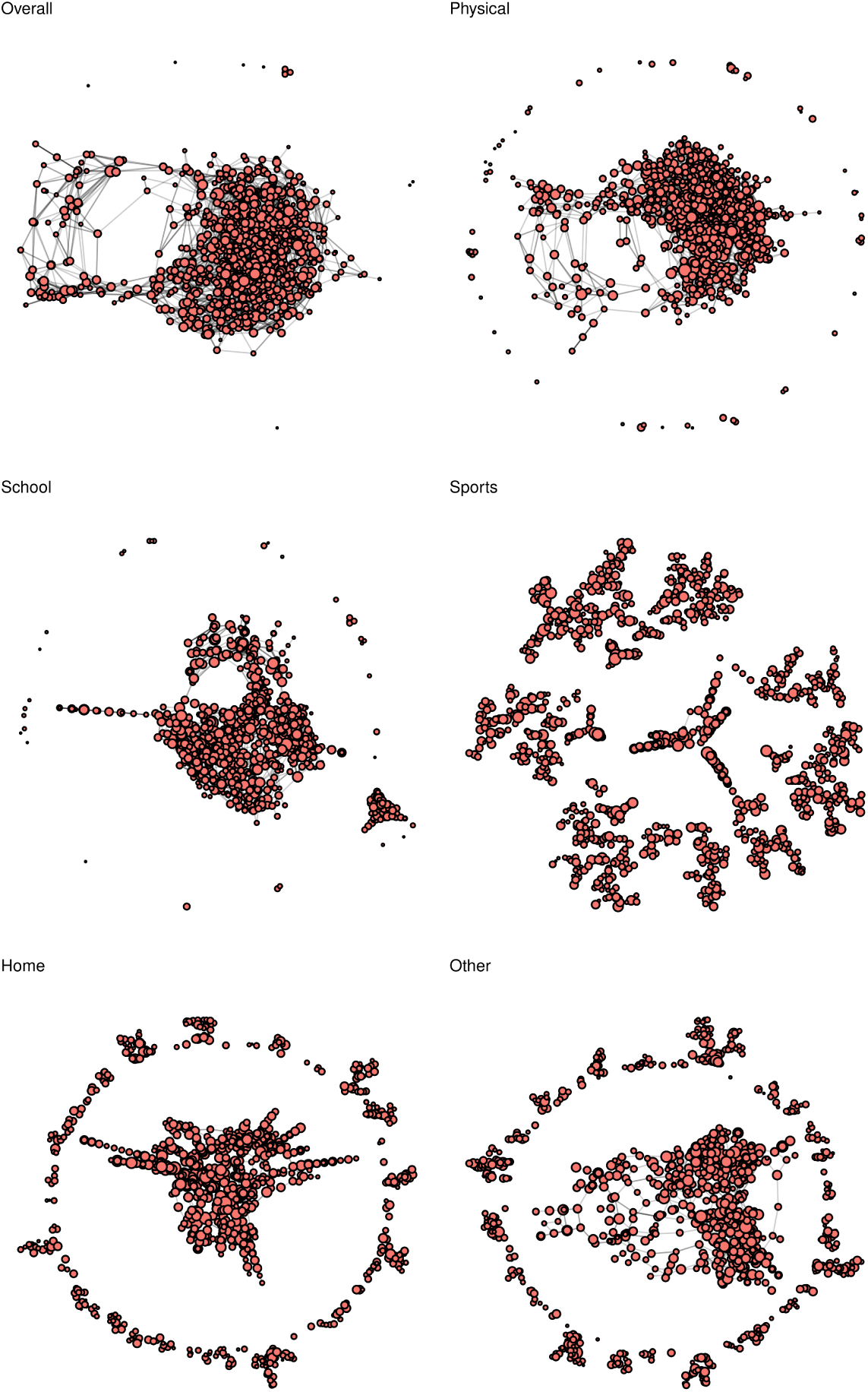
Overview of all friendship networks. The figure is reproduced from “Social network analysis of Staphylococcus aureus carriage in a general youth population” with permissions. DOI: https://doi.org/10.1016/j.ijid.2022.08.018

### 11.2 Network relevance overview

**Figure 7:**
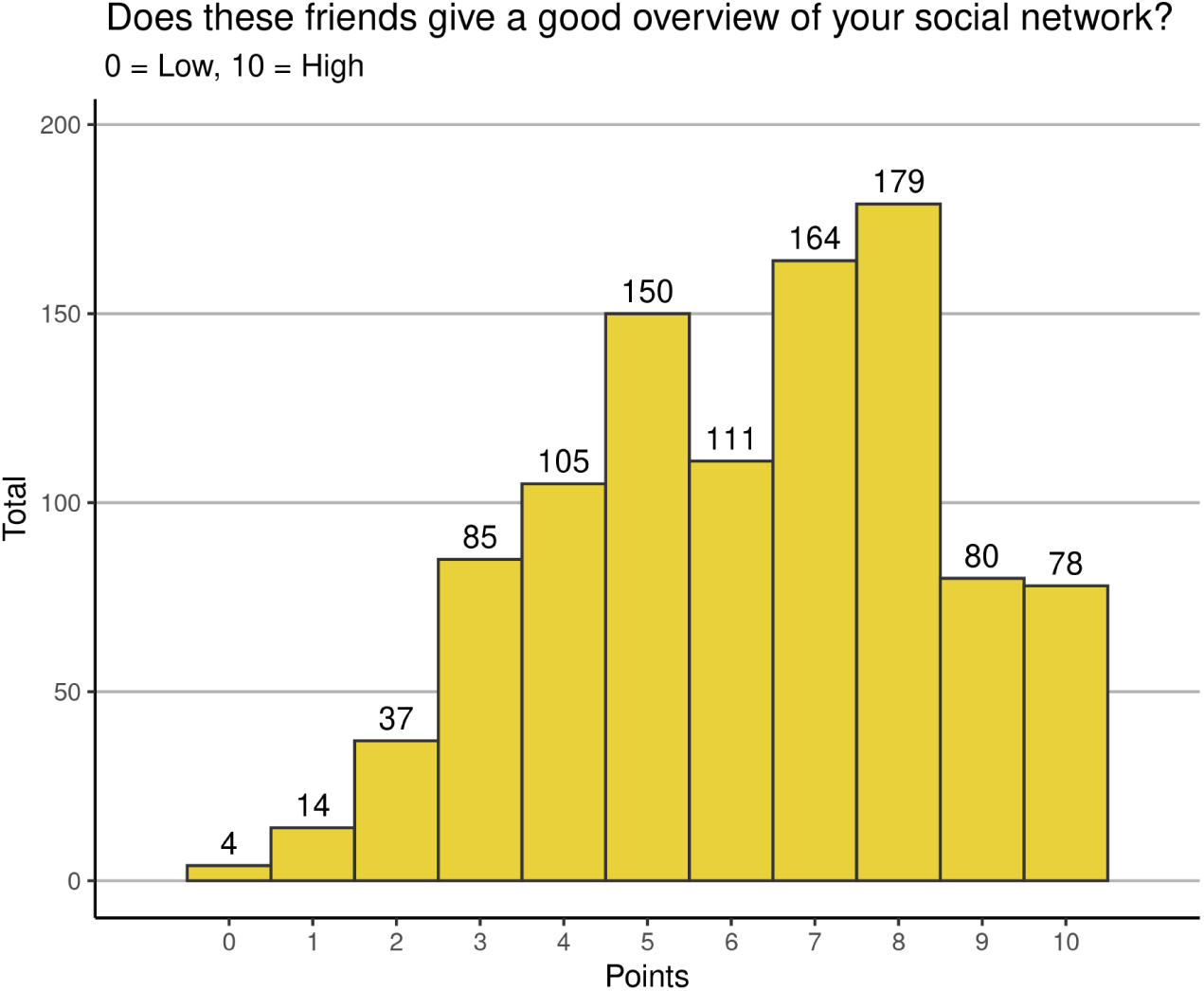
Histogram with how good is the self-reported network (0 - 10, x-axis) by each student (y-axis). The figure is reproduced from “Social network analysis of Staphylococcus aureus carriage in a general youth population” with permissions. DOI: https://doi.org/10.1016/j.ijid.2022.08.018

### 11.3 Network relevance by high school

**Figure 8:**
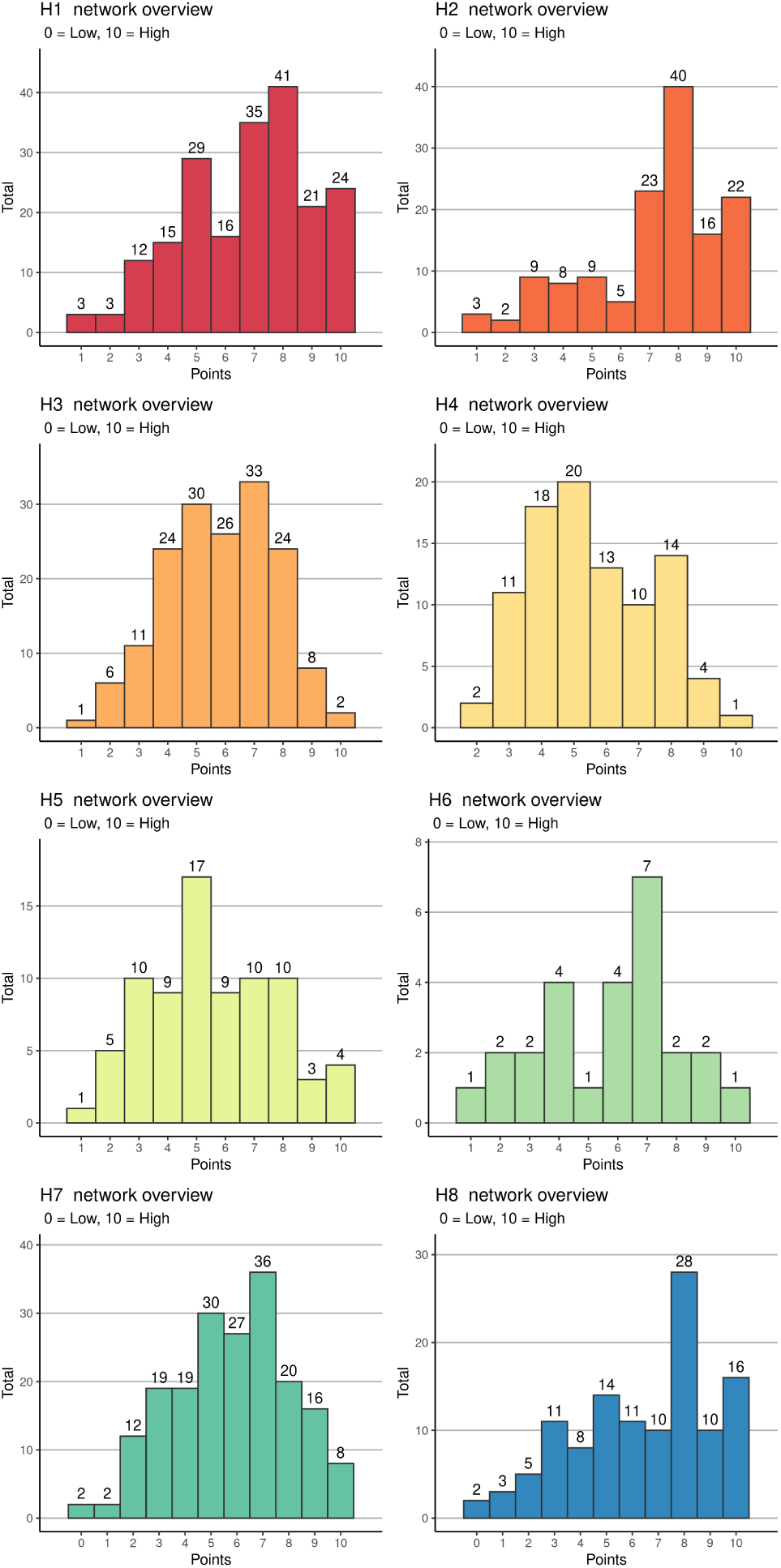
For each of the high schools (H1 to H8), histogram with how good is the self-reported network (0 - 10, x-axis) by each student (y-axis)

### 11.4 Network relevance table

**Table 4:** Highschools and friendship overview. The first row represents all high schools combined. H1 to H8 rows represent each high school separately. Homophily represents how many students of this school form friendships with a student of the same school. Frequency is the relative frequency of students in each high school. Significance is the p-value of a two-sided binomial test of relationships within the same high school, total relationships, and relative frequency of students in each high school. Average and Median are the values of how good is the self-reported network (0 - 10) by each student in each high school.

| High School | Homophily | Frequency | Significance | Average | Median |
| --- | --- | --- | --- | --- | --- |
| All | 0.87 |  |  | 6.22 | 6 |
| H1 | 0.76 | 0.2 | **** | 6.77 | 7 |
| H2 | 0.78 | 0.14 | **** | 7.2 | 8 |
| H3 | 0.76 | 0.16 | **** | 5.84 | 6 |
| H4 | 0.77 | 0.09 | **** | 5.54 | 5 |
| H5 | 0.96 | 0.08 | **** | 5.55 | 5 |
| H6 | 0.76 | 0.03 | **** | 5.73 | 6 |
| H7 | 0.79 | 0.18 | **** | 5.8 | 6 |
| H8 | 0.72 | 0.12 | **** | 6.42 | 7 |

### 11.5 Non-solarium networks by high school

**Figure 9:**
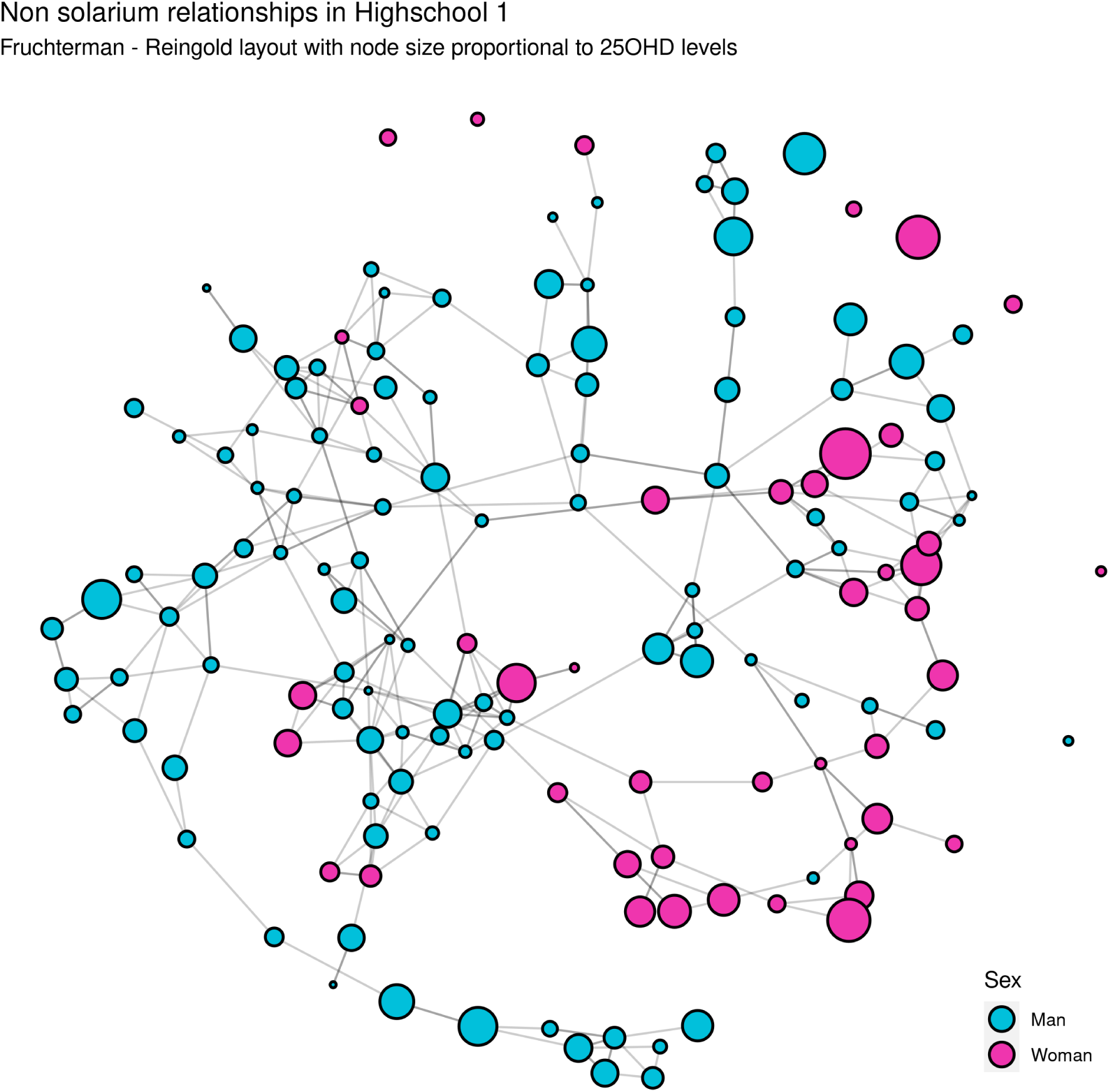
Relationships in H1 for non-solarium users highlighted by sex. Node size is proportional to the 25OHD level. Layout of the nodes using Fruchterman - Reingold. A total of 150 students and 255 undirected relationships are displayed.

**Figure 10:**
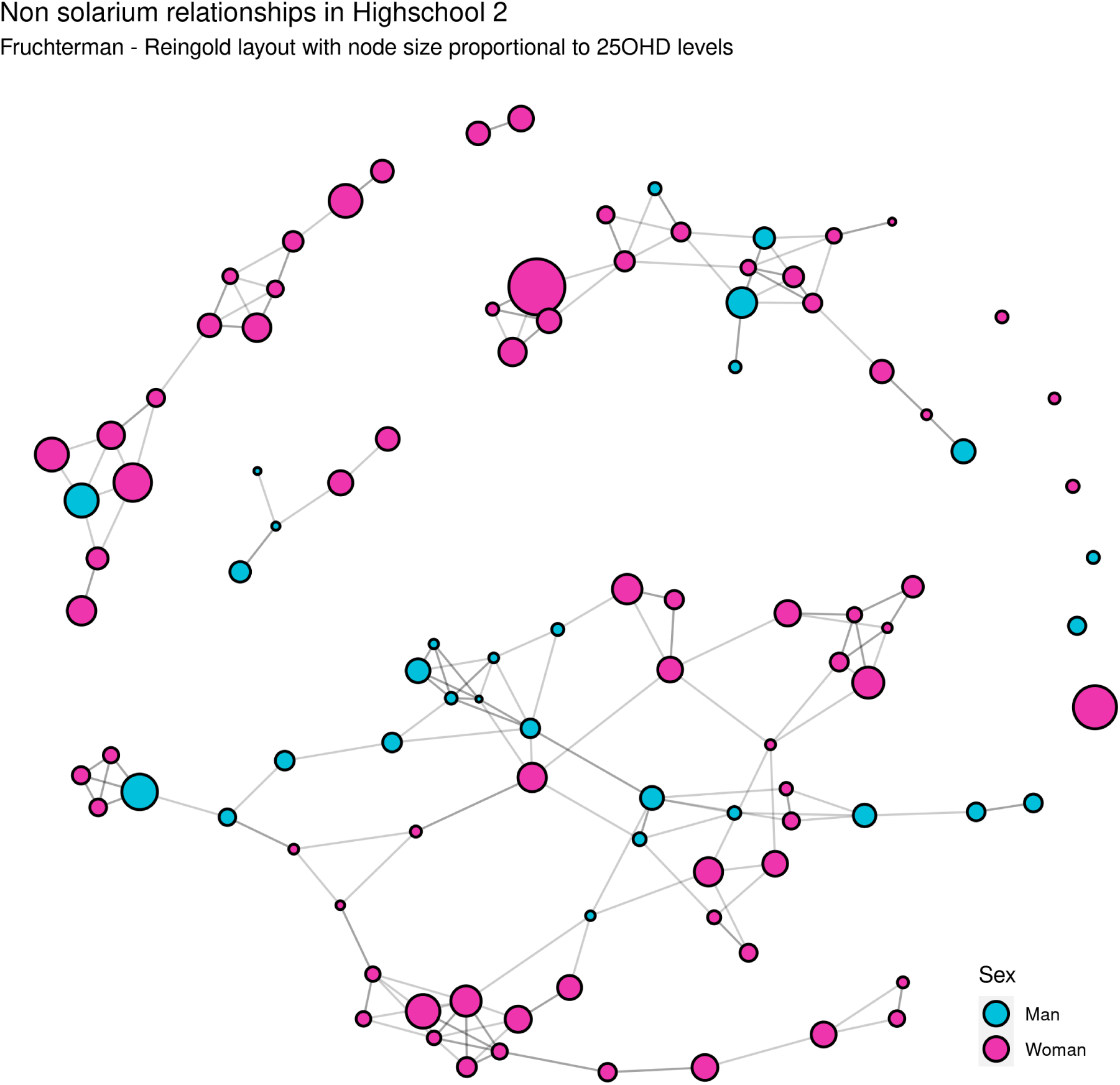
Relationships in H2 for non-solarium users highlighted by sex. Node size is proportional to the 25OHD level. Layout of the nodes using Fruchterman - Reingold. A total of 101 students and 157 undirected relationships are displayed.

**Figure 11:**
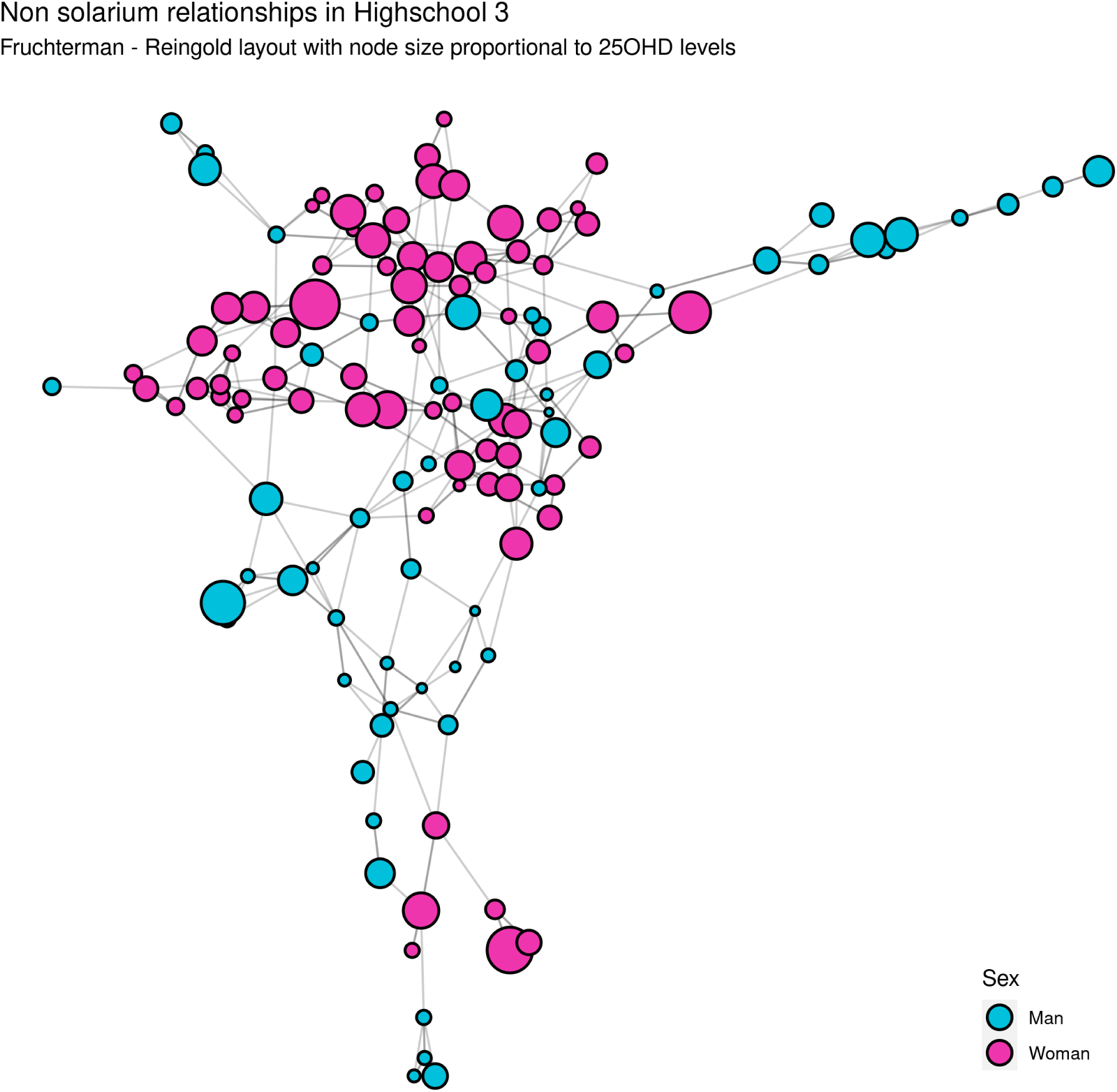
Relationships in H3 for non-solarium users highlighted by sex. Node size is proportional to the 25OHD level. Layout of the nodes using Fruchterman - Reingold. A total of 129 students and 247 undirected relationships are displayed.

**Figure 12:**
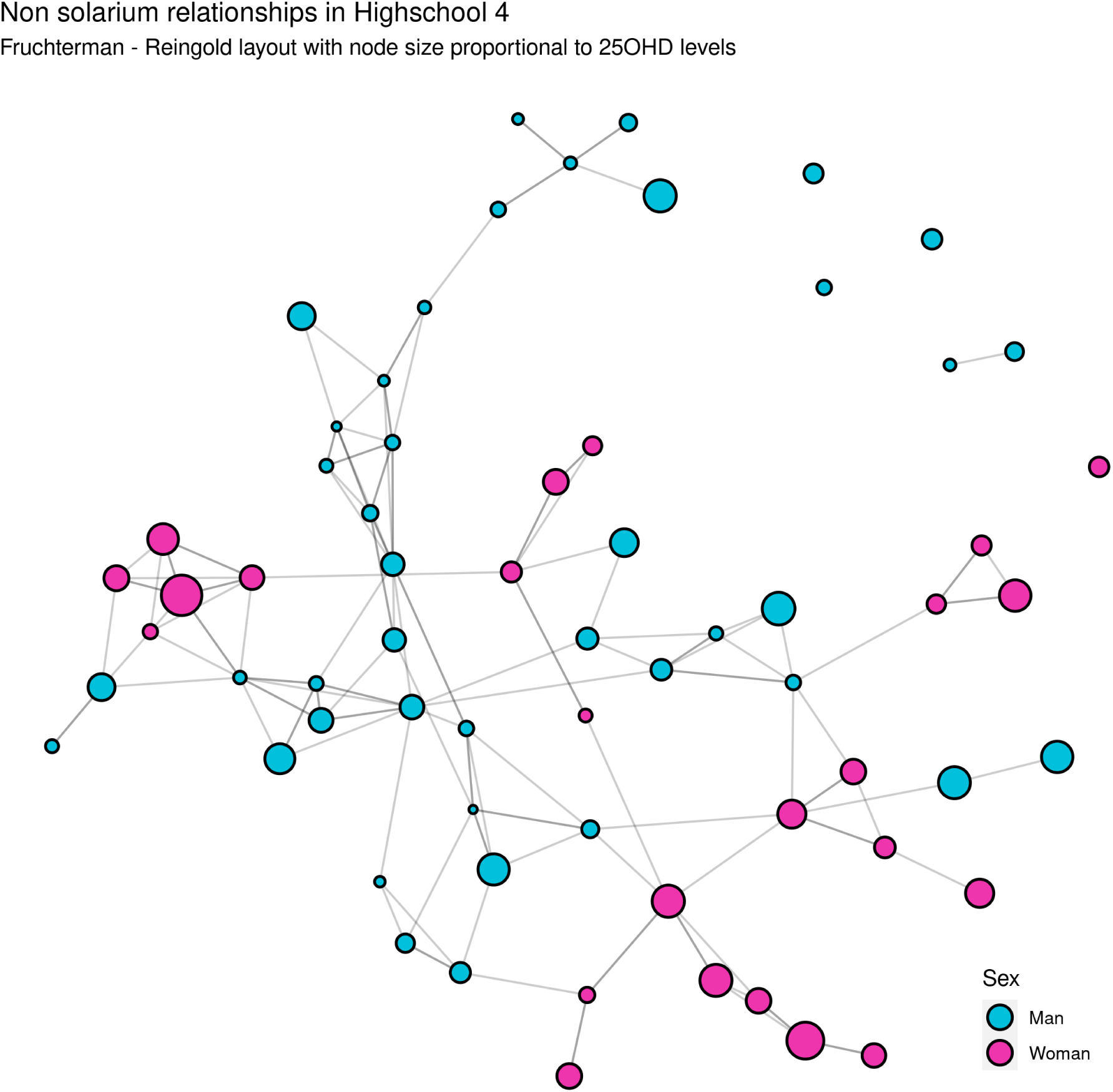
Relationships in H4 for non-solarium users highlighted by sex. Node size is proportional to the 25OHD level. Layout of the nodes using Fruchterman - Reingold. A total of 65 students and 110 undirected relationships are displayed.

**Figure 13:**
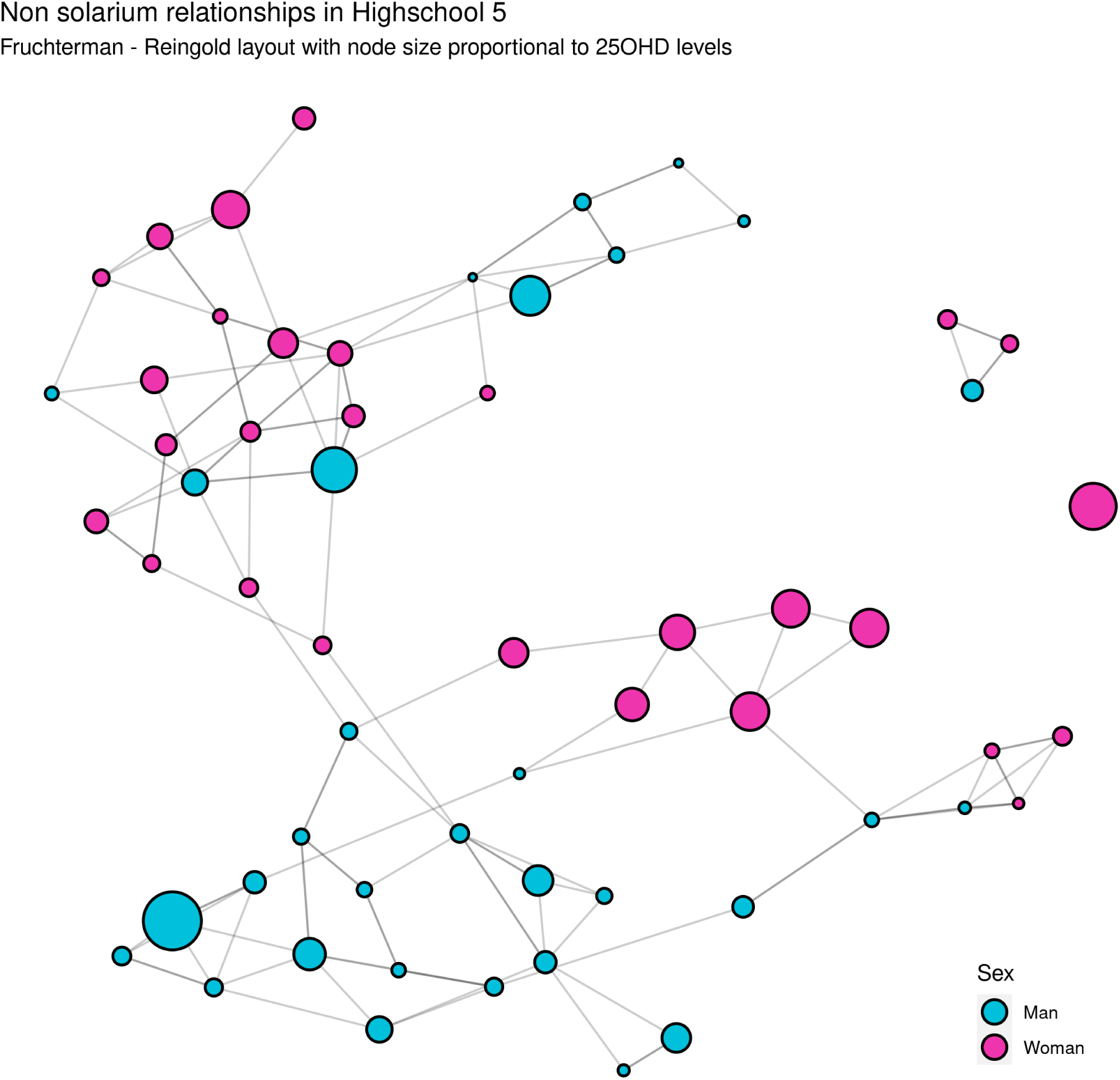
Relationships in H5 for non-solarium users highlighted by sex. Node size is proportional to the 25OHD level. Layout of the nodes using Fruchterman - Reingold. A total of 59 students and 102 undirected relationships are displayed.

**Figure 14:**
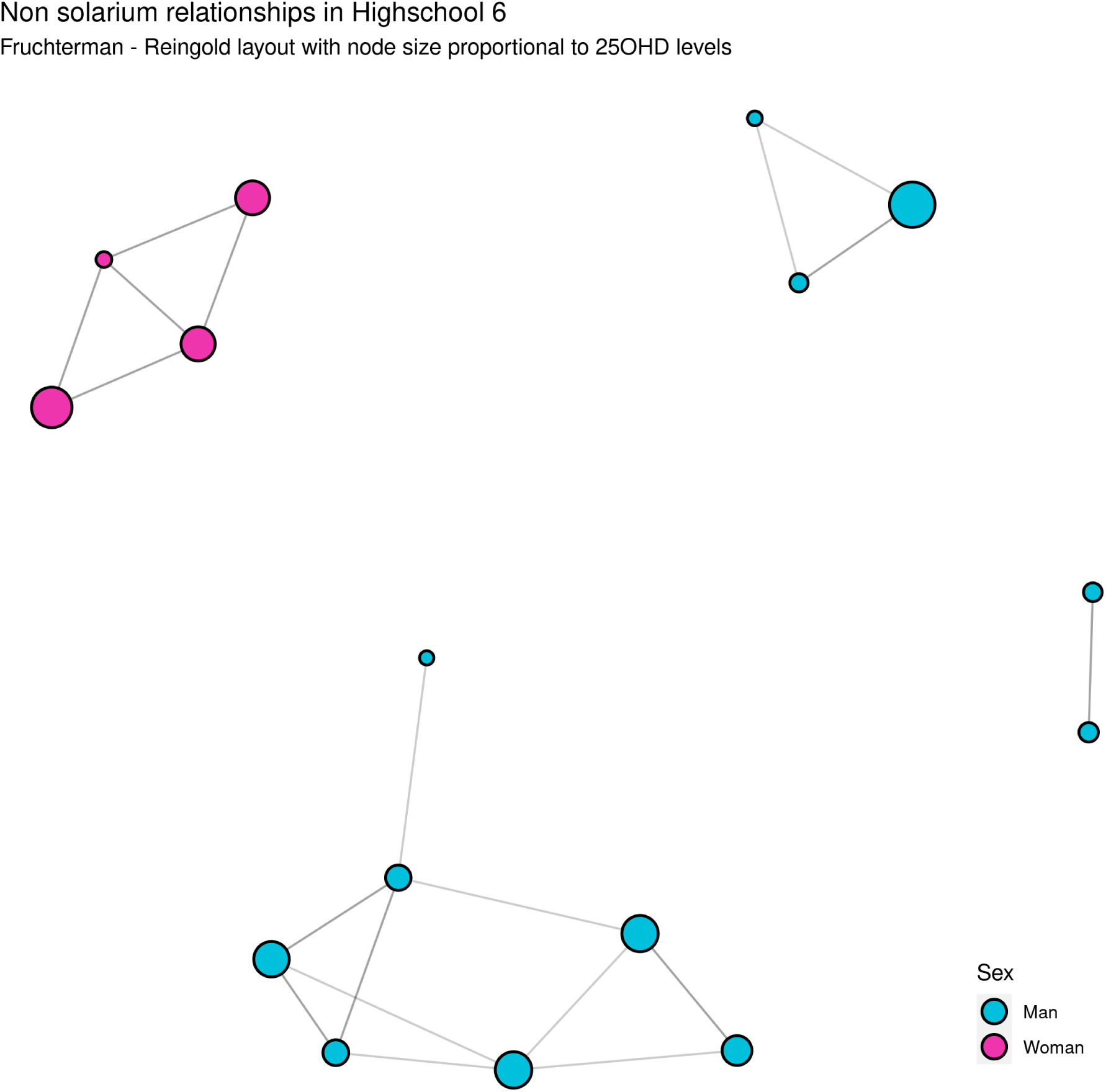
Relationships in H7 for non-solarium users highlighted by sex. Node size is proportional to the 25OHD level. Layout of the nodes using Fruchterman - Reingold. A total of 16 students and 19 undirected relationships are displayed.

**Figure 15:**
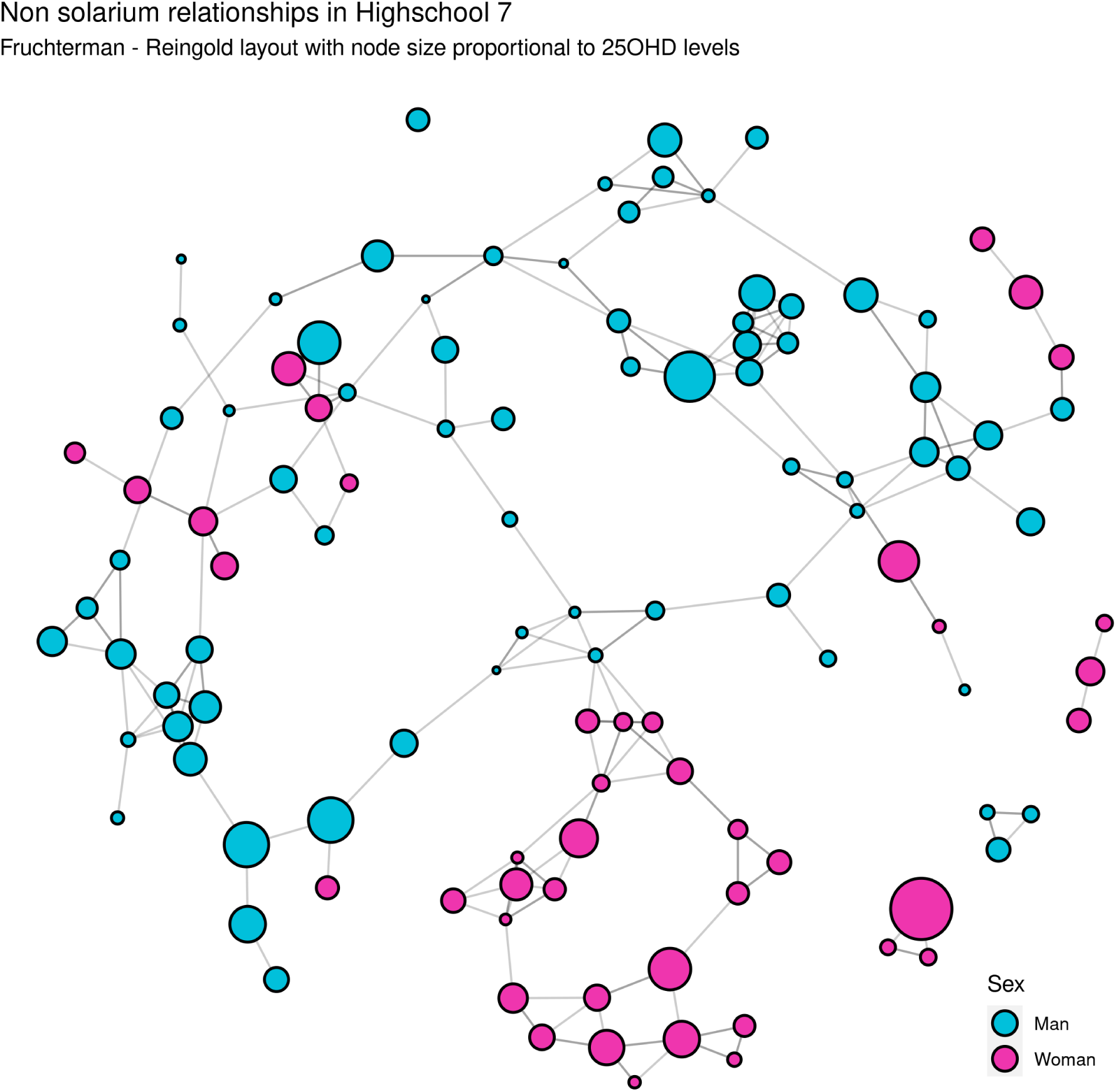
Relationships in H7 for non-solarium users highlighted by sex. Node size is proportional to the 25OHD level. Layout of the nodes using Fruchterman - Reingold. A total of 113 students and 180 undirected relationships are displayed.

**Figure 16:**
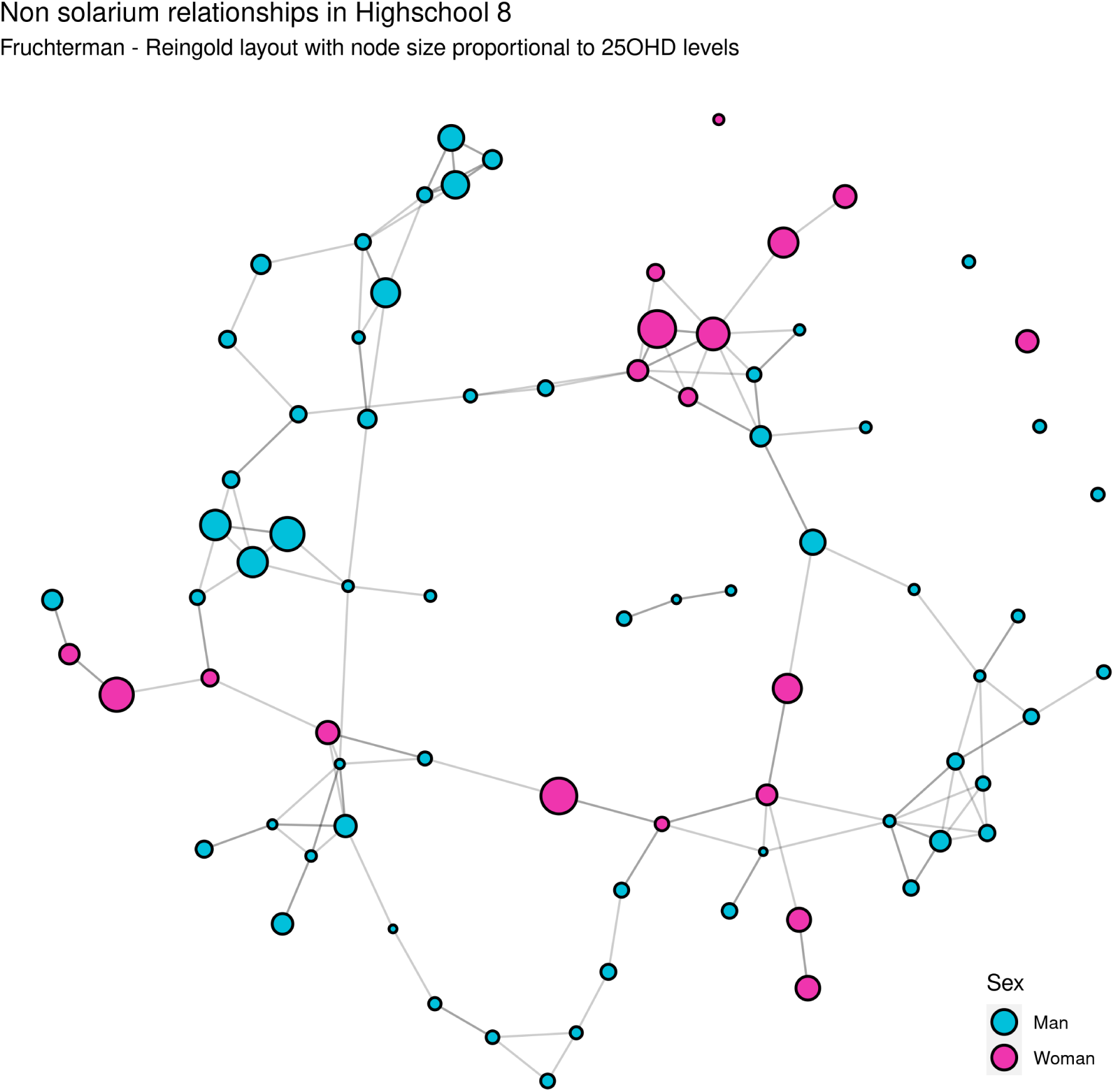
Relationships in H8 for non-solarium users highlighted by sex. Node size is proportional to the 25OHD level. Layout of the nodes using Fruchterman - Reingold. A total of 78 students and 112 undirected relationships are displayed.

## 12 Other social influences

### 12.1 Non-solarium simulations by high school

**Table 5:** Simulations performed for each high school independently for non-solarium users.

|  | Relationships |  | Simulations statistics |  |  |  |  |  |  |  |
| --- | --- | --- | --- | --- | --- | --- | --- | --- | --- | --- |
| Highschool | Total | Equal | MIN | Q1 | Median | Average | Q3 | MAX | SD | P-value |
| H1 | 341 | 223 | 151 | 169 | 175 | 177.16 | 186 | 211 | 12.45 | *** |
| H2 | 221 | 137 | 92 | 109 | 116.5 | 116.2 | 123 | 154 | 10.26 | * |
| H3 | 348 | 212 | 156 | 170 | 181 | 180.99 | 189 | 227 | 13.88 | * |
| H4 | 153 | 101 | 57 | 74 | 81 | 81.24 | 88 | 104 | 9.75 | * |
| H5 | 135 | 88 | 47 | 65 | 71 | 70.22 | 76 | 95 | 8.35 | * |
| H6 | 30 | 20 | 9 | 13 | 16 | 15.96 | 19 | 23 | 3.42 | ns |
| H7 | 251 | 141 | 102 | 123 | 132 | 131.05 | 138 | 159 | 10.52 | ns |
| H8 | 152 | 114 | 63 | 72 | 78 | 78.46 | 85 | 98 | 8.28 | **** |

### 12.2 25OHD and Friends’ average 25OHD by high school

**Figure 17:**
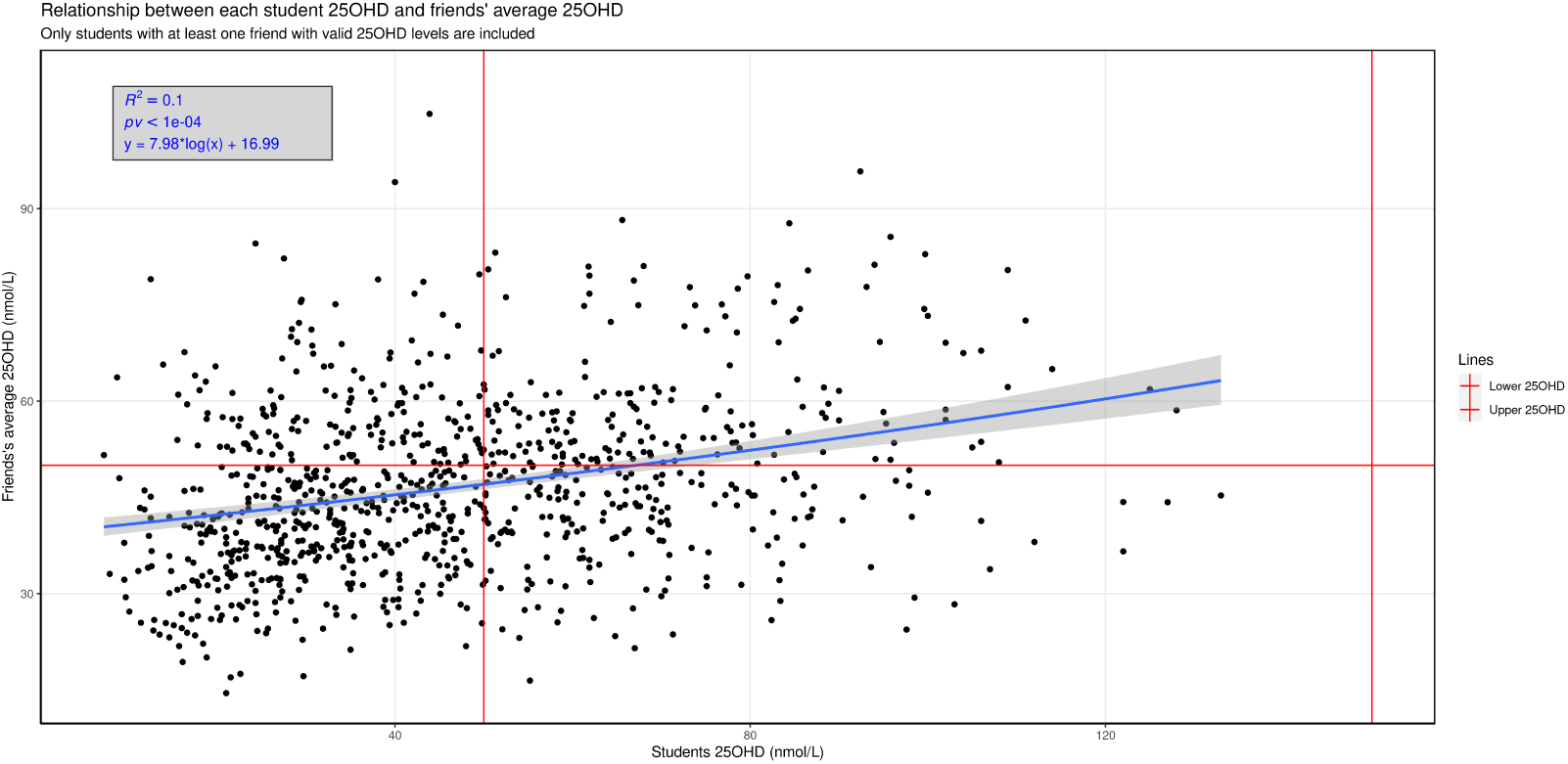
Relationship between each student 25OHD levels (X-axis) in comparison with the students’ friends average 25OHD levels (Y-axis). Only students with a valid 25OHD value with at least one friend who also has a valid 25OHD value are included (n=930)

**Figure 18:**
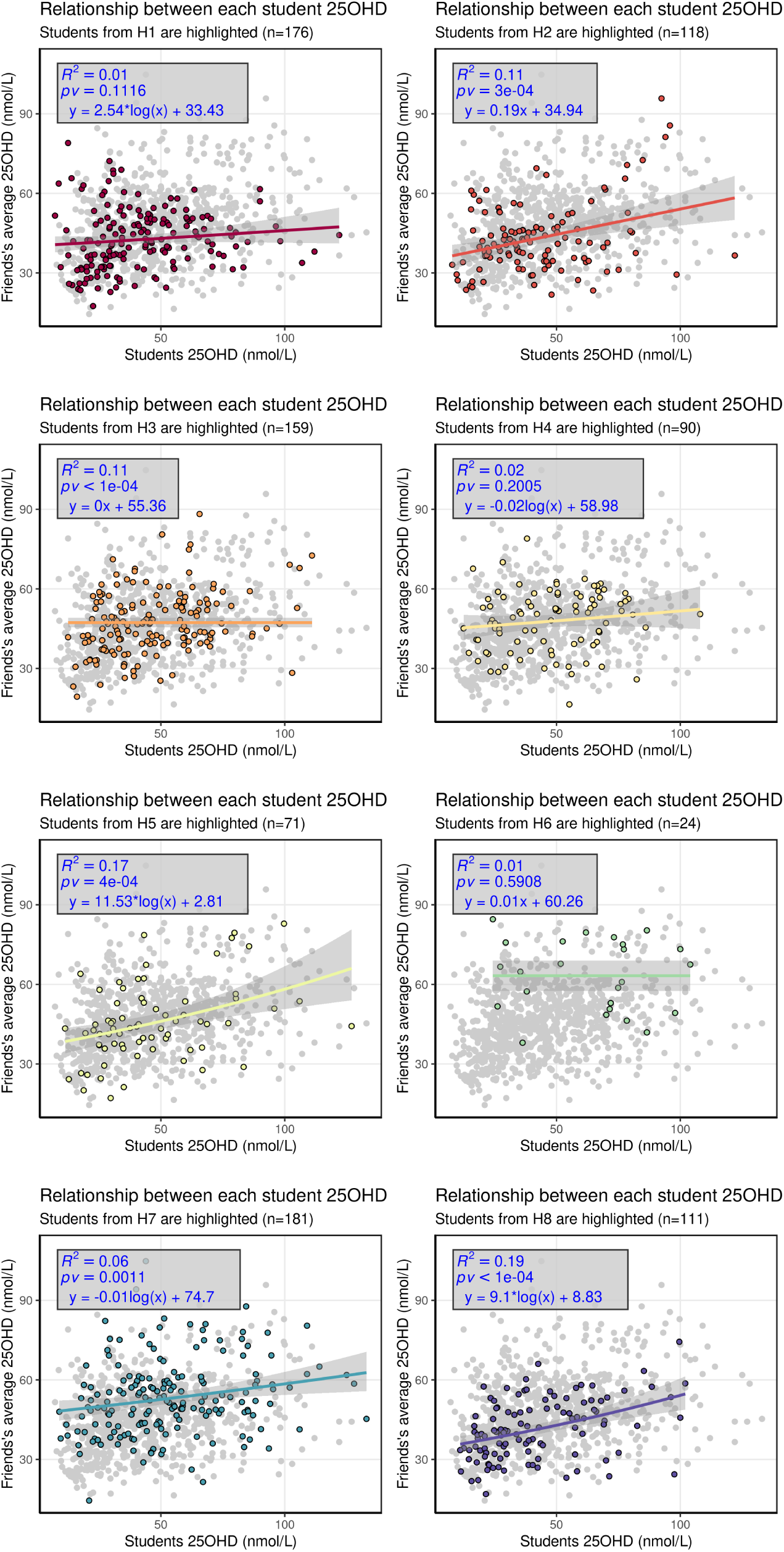
Relationship between each student 25OHD levels in comparison with the students’ friends average 25OHD levels, for each high school. Students from other high schools are grayed out.

## 13 Ethnicity

**Table 6:** Absolute frequency for each ethnicity sorted by frequency.

| Modality | Count | Relative | Cumulative |
| --- | --- | --- | --- |
| Norwegian | 900 | 0.867 | 0.867 |
| Norwegian-Sami | 27 | 0.026 | 0.893 |
| Didn't Answer | 20 | 0.019 | 0.912 |
| Norwegian-Kven | 10 | 0.01 | 0.922 |
| Sami | 7 | 0.007 | 0.929 |
| Norwegian-Swedish | 7 | 0.007 | 0.935 |
| Russian | 4 | 0.004 | 0.939 |
| Norwegian-Sami-Kven | 4 | 0.004 | 0.943 |
| African | 3 | 0.003 | 0.946 |
| Afghan | 3 | 0.003 | 0.949 |
| Swedish | 3 | 0.003 | 0.952 |
| Norwegian-Somalian | 3 | 0.003 | 0.955 |
| Norwegian-Spanish | 3 | 0.003 | 0.958 |
| Norwegian-Danish | 3 | 0.003 | 0.961 |
| Other | 2 | 0.002 | 0.962 |
| Eritrean | 2 | 0.002 | 0.964 |
| Thai | 2 | 0.002 | 0.966 |
| German | 2 | 0.002 | 0.968 |
| Polish | 2 | 0.002 | 0.97 |
| Kven | 2 | 0.002 | 0.972 |
| Norwegian-Russian | 2 | 0.002 | 0.974 |
| Canadian | 1 | 0.001 | 0.975 |
| Tamil | 1 | 0.001 | 0.976 |
| Palestinian | 1 | 0.001 | 0.977 |
| Italian | 1 | 0.001 | 0.978 |
| Bulgarian | 1 | 0.001 | 0.979 |
| Belgian | 1 | 0.001 | 0.98 |
| Dutch | 1 | 0.001 | 0.981 |
| Norwegian-Other | 1 | 0.001 | 0.982 |
| Norwegian-Colombian | 1 | 0.001 | 0.983 |
| Norwegian-Brasilian | 1 | 0.001 | 0.984 |
| Norwegian-Canadian | 1 | 0.001 | 0.985 |
| Norwegian-Ghanaian | 1 | 0.001 | 0.986 |
| Norwegian-Gambian | 1 | 0.001 | 0.987 |
| Norwegian-African | 1 | 0.001 | 0.987 |
| Norwegian-Philipine | 1 | 0.001 | 0.988 |
| Norwegian-Tamil | 1 | 0.001 | 0.989 |
| Norwegian-Thai | 1 | 0.001 | 0.99 |
| Norwegian-Chinese | 1 | 0.001 | 0.991 |
| Norwegian-Turkish | 1 | 0.001 | 0.992 |
| Norwegian-Portuguese | 1 | 0.001 | 0.993 |
| Norwegian-Italian | 1 | 0.001 | 0.994 |
| Norwegian-Belgian | 1 | 0.001 | 0.995 |
| Norwegian-German | 1 | 0.001 | 0.996 |
| Norwegian-Finnish | 1 | 0.001 | 0.997 |
| Norwegian-Swedish-Dutch | 1 | 0.001 | 0.998 |
| Norwegian-Sami-Icelandic | 1 | 0.001 | 0.999 |
| Norwegian-Sami-Swedish | 1 | 0.001 | 1 |

**Table 7:** Xi-square test skin group and high-school.

| 0.5 | Fair | Dark | Total | Freq |
| --- | --- | --- | --- | --- |
| H1 | 190/192 | 7/4 | 197 | 0.19 |
| H2 | 135/136 | 5/3 | 140 | 0.14 |
| H3 | 164/163 | 3/3 | 167 | 0.16 |
| H4 | 95/94 | 2/2 | 97 | 0.1 |
| H5 | 81/79 | 0/1 | 81 | 0.08 |
| H6 | 26/25 | 0/0 | 26 | 0.03 |
| H7 | 188/186 | 3/4 | 191 | 0.19 |
| H8 | 117/116 | 2/2 | 119 | 0.12 |
| Total | 996 | 22 | 1018 |  |
| Freq | 0.98 | 0.02 |  | 1 |

## 14 Non solarium distributions by high schools

### 14.1 Sex

**Table 8:** Xi2 table with respect to sex and high school.

| 7e-13 | Man | Woman | Total | Freq |
| --- | --- | --- | --- | --- |
| H1 | 107/85 ++ | 43/64 — | 150 | 0.21 |
| H2 | 29/57 — | 72/43 +++++ | 101 | 0.14 |
| H3 | 56/73 - | 73/55 ++ | 129 | 0.18 |
| H4 | 41/37 | 24/27 | 65 | 0.09 |
| H5 | 31/33 | 28/25 | 59 | 0.08 |
| H6 | 12/9 | 4/6 | 16 | 0.02 |
| H7 | 71/64 | 42/48 | 113 | 0.16 |
| H8 | 59/44 ++ | 19/33 — | 78 | 0.11 |
| Total | 406 | 305 | 711 |  |
| Freq | 0.57 | 0.43 |  | 1 |

### 14.2 BMI

**Table 9:** Xi2 table with respect to BMI and high school.

| 9e-04 | Underweight | Healthy | Overweight | Obese | Total | Freq |
| --- | --- | --- | --- | --- | --- | --- |
| H1 | 16/16 | 91/100 | 26/20 | 16/11 | 149 | 0.21 |
| H2 | 14/11 | 57/67 | 16/13 | 13/7 + | 100 | 0.14 |
| H3 | 13/14 | 96/86 | 15/17 | 5/10 - | 129 | 0.18 |
| H4 | 12/7 + | 39/43 | 9/8 | 5/5 | 65 | 0.09 |
| H5 | 4/6 | 40/39 | 6/8 | 9/4 ++ | 59 | 0.08 |
| H6 | 0/1 | 16/10 | 0/2 | 0/1 | 16 | 0.02 |
| H7 | 15/12 | 88/75 | 8/15 - | 2/8 — | 113 | 0.16 |
| H8 | 6/8 | 49/52 | 17/10 ++ | 6/6 | 78 | 0.11 |
| Total | 80 | 476 | 97 | 56 | 709 |  |
| Freq | 0.11 | 0.67 | 0.14 | 0.08 |  | 1 |

### 14.3 Alcohol

**Table 10:** Xi2 table with respect to alcohol and high school for non-solarium users.

| 0.001 | Never | Once per month or less | Twice of more per month | Total | Freq |
| --- | --- | --- | --- | --- | --- |
| H1 | 44/45 | 60/56 | 37/38 | 141 | 0.2 |
| H2 | 36/32 | 38/40 | 26/27 | 100 | 0.14 |
| H3 | 40/41 | 61/52 | 28/35 | 129 | 0.19 |
| H4 | 25/20 | 23/25 | 16/17 | 64 | 0.09 |
| H5 | 17/18 | 17/22 | 23/15 + | 57 | 0.08 |
| H6 | 11/5 ++ | 5/6 | 0/4 - | 16 | 0.02 |
| H7 | 38/36 | 48/45 | 27/30 | 113 | 0.16 |
| H8 | 14/24 - | 29/31 | 34/21 +++ | 77 | 0.11 |
| Total | 225 | 281 | 191 | 697 |  |
| Freq | 0.32 | 0.4 | 0.27 |  | 1 |

### 14.4 Smoke

**Table 11:**
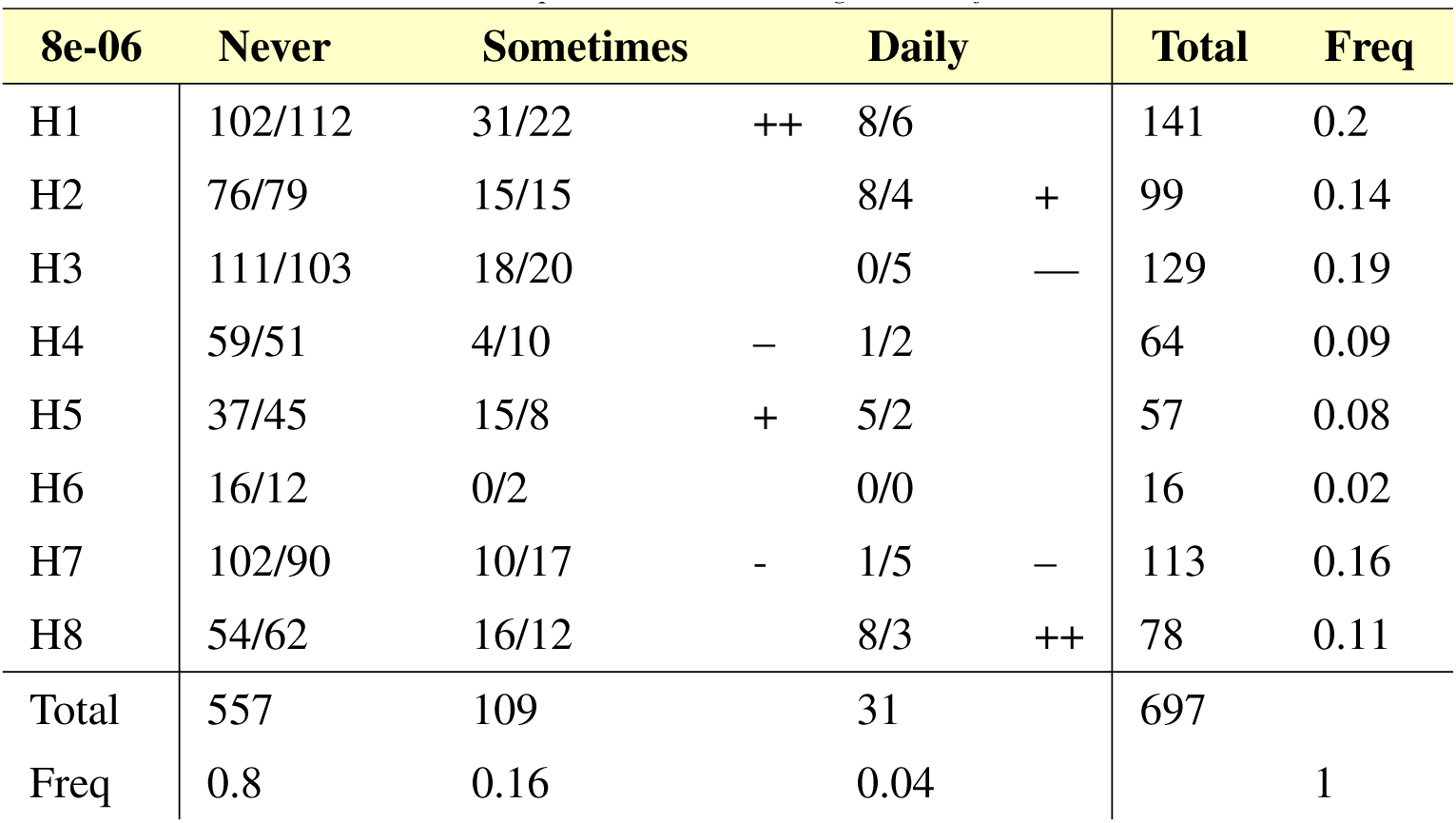
Xi2 table with respect to smoke and high school for non-solarium users.

### 14.5 Snuff

**Table 12:**
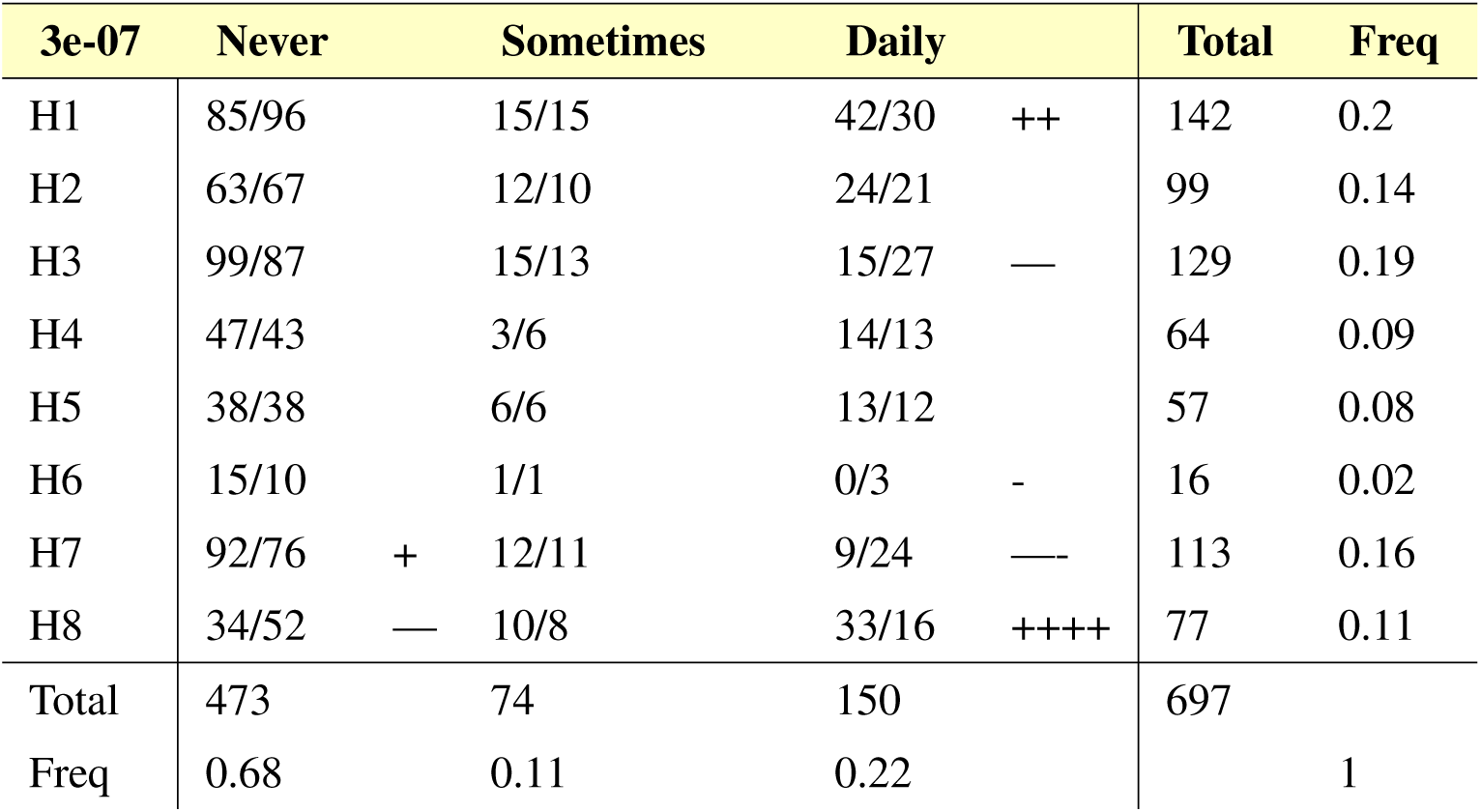
Xi2 table with respect to snuff and high school for non-solarium users.

### 14.6 Sports

**Table 13:** Xi2 table with respect to sport and high school for non-solarium users.

| 9e-34 | None | Light | Medium | Hard | Total | Freq |
| --- | --- | --- | --- | --- | --- | --- |
| H1 | 44/36 | 63/47 ++ | 30/34 | 5/23 — | 142 | 0.2 |
| H2 | 38/25 ++ | 40/33 | 14/24 – | 8/16 – | 100 | 0.14 |
| H3 | 24/33 - | 44/42 | 42/31 ++ | 19/21 | 129 | 0.18 |
| H4 | 16/16 | 14/21 | 21/15 | 13/10 | 64 | 0.09 |
| H5 | 15/14 | 19/18 | 21/13 + | 2/9 — | 57 | 0.08 |
| H6 | 0/4 – | 0/5 — | 0/3 – | 16/2 +++++ | 16 | 0.02 |
| H7 | 12/29 — | 22/37 — | 33/27 | 46/18 +++++ | 113 | 0.16 |
| H8 | 32/20 +++ | 30/25 | 10/19 – | 6/12 – | 78 | 0.11 |
| Total | 181 | 232 | 171 | 115 | 699 |  |
| Freq | 0.26 | 0.33 | 0.24 | 0.16 |  | 1 |

### 14.7 Sunbathing

**Table 14:** Xi-square test for sunbathing and high school for non-solarium users.

| 4e-07 | Yes | No | Total | Freq |
| --- | --- | --- | --- | --- |
| H1 | 25/10 +++++ | 124/138 | 149 | 0.21 |
| H2 | 5/7 | 96/93 | 101 | 0.14 |
| H3 | 5/9 | 124/119 | 129 | 0.18 |
| H4 | 1/4 - | 64/60 | 65 | 0.09 |
| H5 | 2/4 | 57/54 | 59 | 0.08 |
| H6 | 5/1 +++ | 11/14 | 16 | 0.02 |
| H7 | 6/8 | 107/104 | 113 | 0.16 |
| H8 | 3/5 | 74/71 | 77 | 0.11 |
| Total | 52 | 657 | 709 |  |
| Freq | 0.07 | 0.93 |  | 1 |

## 15 Other figures

### 15.1 PTH predictors

**Figure 19:**
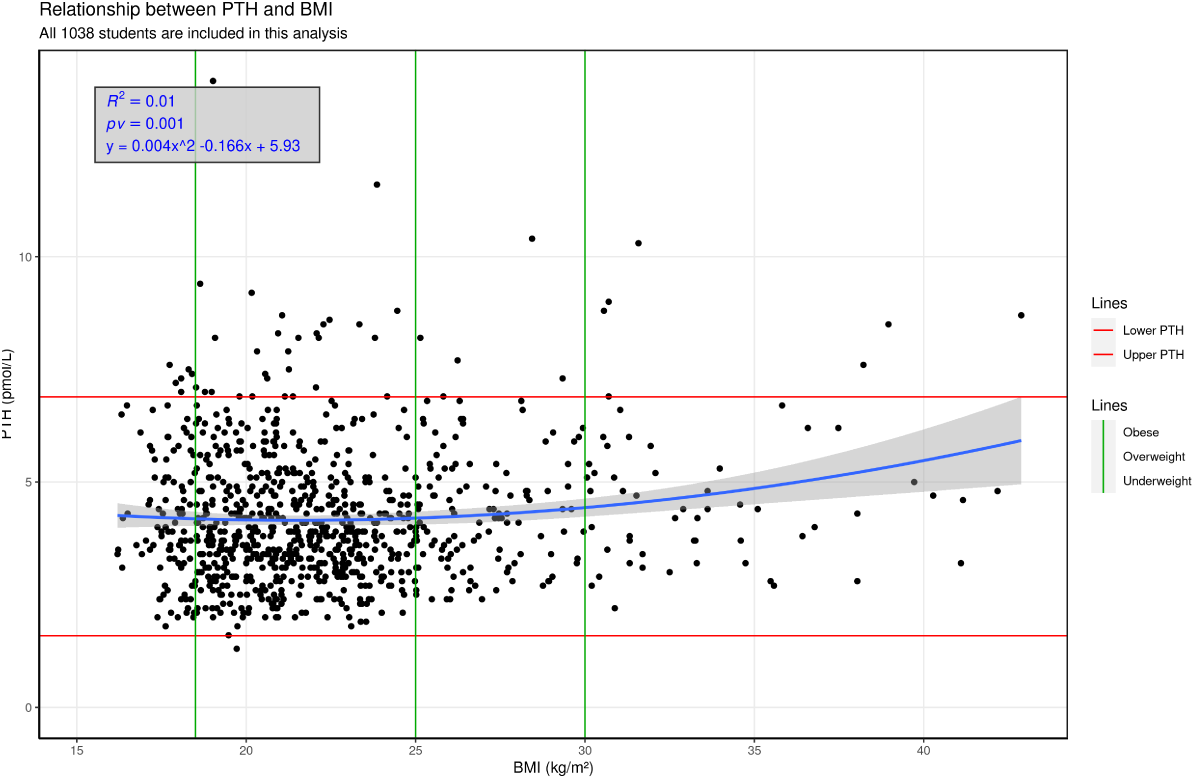
Relationship between PTH and BMI. All the students are included in this analysis. Vertical lines represent the threshold between BMI categories. Horizontal lines represent the healthy boundaries of PTH levels.

**Figure 20:**
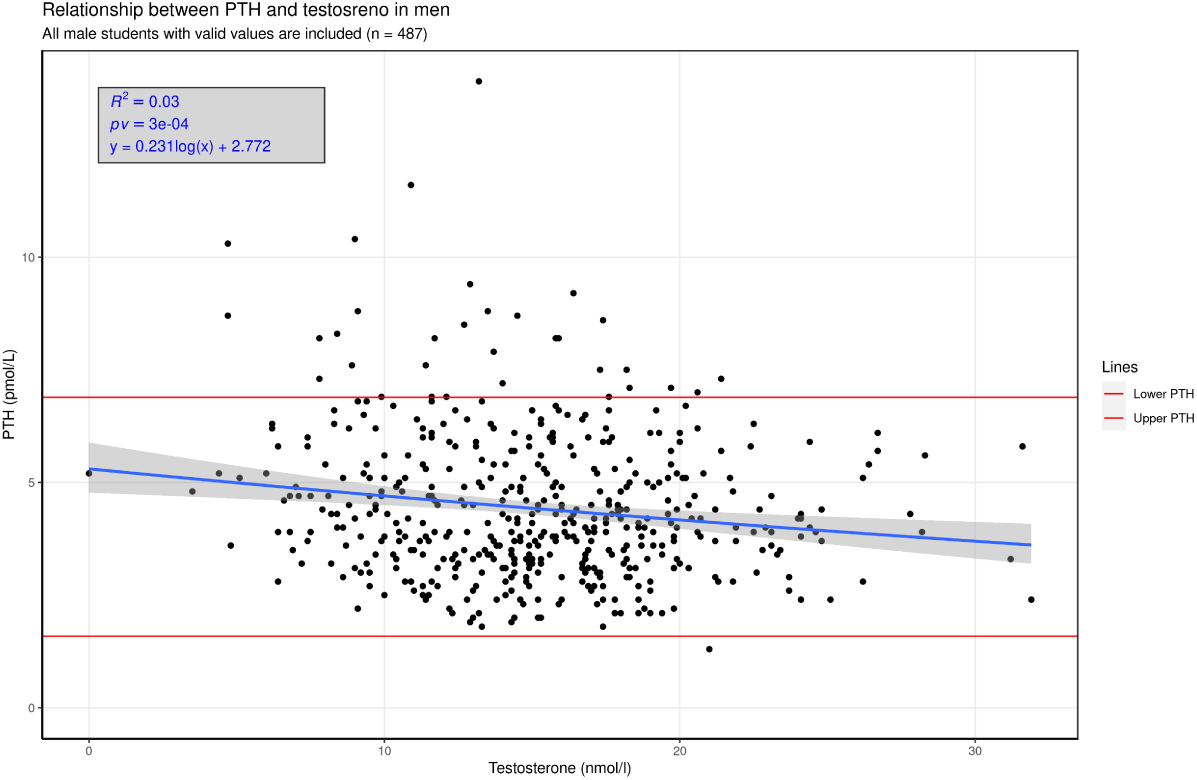
Relationship between PTH and testosterone in men. All the students are included in this analysis. Horizontal lines represent the healthy boundaries of PTH levels.

**Figure 21:**
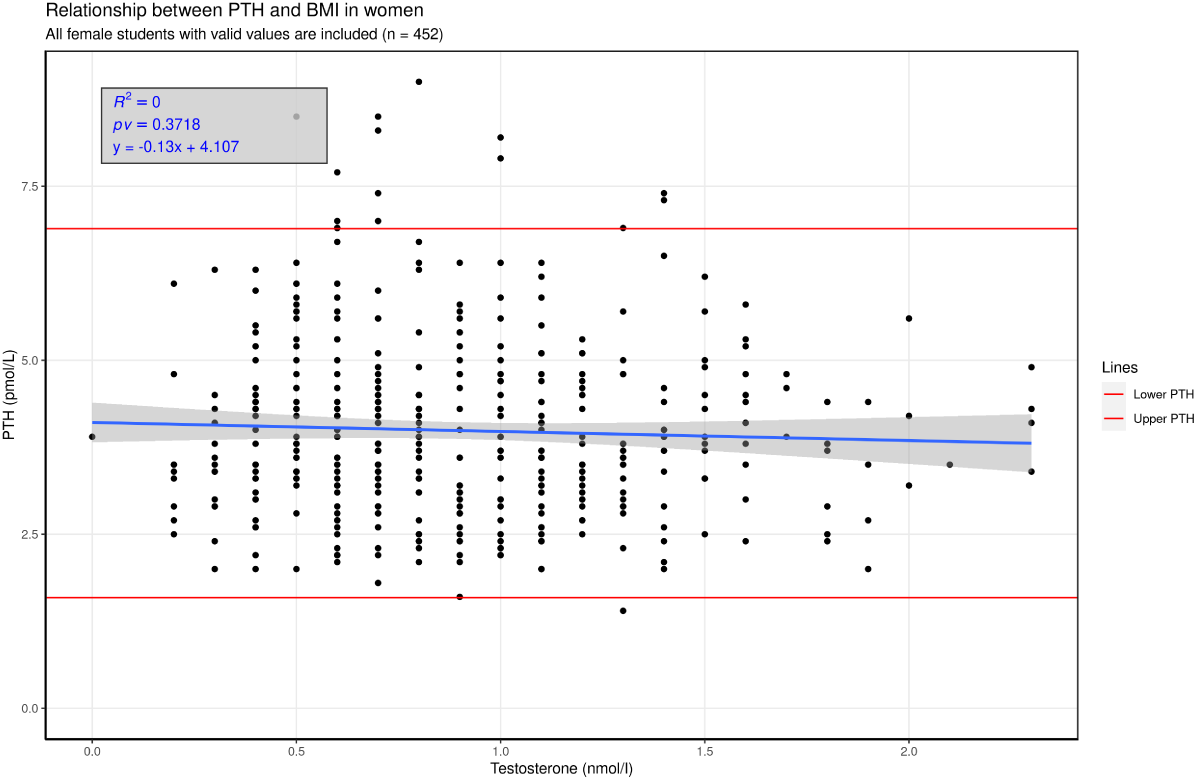
Relationship between PTH and testosterone in women. All the students are included in this analysis. Horizontal lines represent the healthy boundaries of PTH levels.

